# Urinary Pesticide Biomarker Associations with Depression and Anxiety Scores from Adolescence into Young Adulthood in the ESPINA cohort

**DOI:** 10.64898/2025.12.10.25341988

**Authors:** Briana N.C. Chronister, Mohammed N. Hussari, Dolores Lopez-Paredes, Harvey Checkoway, Jose Suarez-Torres, Sheila Gahagan, Asa Bradman, Danilo Martinez, Suzi Hong, Raeanne C. Moore, Jose R. Suarez-Lopez

**Affiliations:** Herbert Wertheim School of Public Health and Human Longevity Science, University of California San Diego, La Jolla, CA, 92093, USA; School of Public Health, San Diego State University, San Diego, CA, 92182, USA; School of Medicine, University of California San Diego (UCSD), La Jolla, CA, USA; Fundación Cimas del Ecuador, Quito, Ecuador; Department of Pediatrics, University of California, San Diego, La Jolla, CA, 92093, USA; Department of Public Health, School of Social Sciences, Humanities, and Arts, University of California, Merced, CA, USA; Department of Psychiatry, University of California San Diego, La Jolla, CA, 92093, USA

## Abstract

**Introduction:** Growing evidence suggests pesticide exposure can affect emotional well-being; limited research exists across adolescence and young adulthood. We examined cross-sectional and longitudinal associations of pesticide biomarkers with anxiety and depression scores.

**Methods:** We analyzed 646 participants from Ecuadorian agricultural communities: 510 in 2016 (ages 11–17y) and 485 in 2022 (17–24y). Twelve urinary insecticide metabolites were measured. Validated questionnaires assessed depression and anxiety. Generalized estimating equations (GEE) estimated associations for continuous scores and logit GEE calculated odds ratios (OR) of elevated symptoms, adjusting for demographic and anthropometric variables.

**Results:** In 2016, an interquartile range higher overall pesticides summed (β=1.01 95%CI:[0.48, 1.53]), organophosphates summed (β=0.99 [0.47, 1.51]), 3,5,6-trichloro-2-pyridinol (TCPy; β=0.97 [0.49, 1.45]), pyrethroids summed (β=0.69 [0.11, 1.27]) and 3-phenoxybenzoic acid (β=0.31 [0.01, 0.61]) were positively associated with depression. Detectable 5-hydroxy imidacloprid (OHIM) doubled elevated depression odds. In 2022, higher sulfoxaflor isomers were protective, but higher clothianidin increased elevated depression odds. Longitudinally, depression was positively associated with overall pesticides summed (β=0.06 [0.02, 0.11]), organophosphates summed (β=0.05 [0.001, 0.10]), TCPy (β=0.06 [0.02, 0.10]), pyrethroids summed and trans-3-(2,2-dichlorovinyl)-2,2-dimethylcyclopropane carboxylic acid (trans-DCCA; β=0.01 [0.003, 0.02]). For anxiety, in 2016 detectable trans-DCCA (OR=2.52 [1.52, 4.16]) was associated with elevated symptoms. In 2022, higher clothianidin increased elevated anxiety odds (OR=1.44 [1.07, 1.93]). Longitudinally, trans-DCCA (OR=1.39 [1.03, 1.87]) and OHIM (OR=1.44 [1.02, 2.04]) increased elevated anxiety odds.

**Conclusions:** Organophosphates, pyrethroids, and neonicotinoids were generally linked to higher depression and anxiety, while sulfoximines and one organophosphate appeared protective. OHIM and trans-DCCA showed consistent adverse associations across years.

## Introduction

In the last 50 years, many new pesticides have been developed and widely used in agricultural and floricultural industries globally, especially in Latin America and developing countries. Their use increases risk of chronic exposure to agricultural communities through drift, residues in homes, and soil, water, or food contamination (Ecobichon, 2001; Ferré et al., 2018; Simcox et al., 1995; Suarez-Lopez et al., 2013). Epidemiological evidence links pesticide-related toxicity to psychiatric disorders, such as depression and anxiety. For example, farmers and other occupationally exposed groups show higher depression and anxiety prevalence than non-occupational populations, particularly following pesticide poisonings (Beseler et al., 2008; Harrison and Mackenzie Ross, 2016, 2016; Sanne et al., 2004; Weisskopf et al., 2013; Wesseling et al., 2010). Similar associations have been observed in para-occupational and non-occupational agricultural populations. For example, female spouses of licensed pesticide applicators had greater odds of experiencing depression (Beseler et al., 2006). In another study, residents of agricultural communities with higher rates of pesticide expenditure were at a greater risk for hospitalization due to mood disorders compared with non-agricultural communities (Meyer et al., 2010). Emerging evidence has linked exposure to insecticides, like organophosphates, pyrethroids, neonicotinoids, and sulfoximines, to adverse mental health outcomes, possibly due to neurotoxicity.

Organophosphates are widely used; their primary mechanism of action is inhibition of acetylcholinesterase (AChE). Acute organophosphate toxicity is well described, with autonomic, cardiopulmonary, neurological, and psychiatric manifestations (King and Aaron, 2015). We have shown that pesticide exposure, indicated by lower AChE activity, may increase depression and anxiety depression and anxiety symptoms among adolescents growing up near pesticide spray sites (Suarez-Lopez et al., 2020, 2019). Long-term organophosphate exposure below acute toxicity thresholds may also result in neurobehavioral alterations (Naughton and Terry, 2018).

Pyrethroids are insecticides with known neurotoxicity due to modification of electrical activity in the nervous system through voltage-dependent sodium channels in excitable membranes (Vijverberg and vanden Bercken, 1990). Pyrethroid exposure, estimated with urinary metabolite 3-PBA, was associated with increased depressive symptoms measured by PHQ-9 in National Health and Nutrition Examination Survey (NHANES) participants (Li et al., 2023).

Neonicotinoids bind to nicotinic acetylcholine receptors (nAChRs), but they exhibit greater toxicity in invertebrates as they have a higher proportion of nAChRs with high affinity for neonicotinoids, whereas vertebrates have fewer nAChRs(Simon-Delso et al., 2015). For this reason, neonicotinoids have been considered safer compared with other pesticide classes in humans, yet growing evidence links their neonicotinoid exposure and adverse health consequences (Zhang and Lu, 2022). In rodents, neonicotinoid exposure at a no-observed-adverse-effect level was found to induce acute anxiety-like behaviors(Hirano et al., 2018; Zhang and Lu, 2022).

Sulfoximine insecticides have a related mechanism of action as neonicotinoids, as they are agonists of nAChRs, but with distinct structures and activity. This difference allows sulfoximines, like sulfoxaflor, to effectively target pests with neonicotinoid resistance(Sparks et al., 2013). To our knowledge, no studies have assessed the effects of sulfoximine exposure on mental health outcomes.

Overall, few studies have examined the longitudinal effects of pesticide exposure on mental health, and none during the transition from adolescence to young adulthood. In a study of Korean adults, those who reported depression were more likely to have reported using pesticides(Koh et al., 2017). Likewise, individuals who reported using pesticides for 20 years or more had over twice the odds of having depression compared to individuals who self-reported not using pesticides (Koh et al., 2017). Among 82 farmers (mean age 49.4 ± 9.1 years), longer farming duration ranging from 10 to 30+ years was positively correlated with depression symptoms. These studies relied on self-reported exposure and exclusively studied adults. As such, the potential impact of objectively measured pesticide exposure on mental health during the critical developmental transition from adolescence to adulthood remains unexamined.

The objective of this study is to identify whether exposure to pesticides, measured by urinary metabolites of organophosphates, neonicotinoids, pyrethroids, and sulfoximines, are associated with depression and anxiety symptoms in the Study of Secondary Exposure to Pesticides Across Childhood, Adolescence and Adulthood (ESPINA), based in agricultural communities in Ecuador.

## Methods

### Participants

Information about ESPINA has been previously published (Kornher et al., 2025; Parajuli et al., 2025; Suarez-Lopez et al., 2017). Briefly, ESPINA is a longitudinal cohort examining effects of pesticide exposure on human health using a life-course approach. In 2008, 313 children (4-9 years) from the agricultural and floricultural canton of Pedro Moncayo, Pichincha province, Ecuador were first examined (Suarez-Lopez et al., 2012). Most were recruited through the 2004 Survey of Access and Demand of Health Services (SADHS), a regionally representative survey developed and administered by Fundación Cimas del Ecuador, the Local Rural Governments of Pedro Moncayo, and community stakeholders. Additional participants were recruited through community announcements and word-of-mouth. Inclusion criteria were: (1) children living with flower workers for ≥1 year; (2) children living without agricultural workers and did not have agricultural pesticides stored in their household. Exclusion criteria were medical conditions preventing study participation.

Since 2008, ESPINA has had thirteen follow-up assessments (8 in-person,5 remote). This study used data from the July to October 2016 (follow-up year [FUY] 8b) and the July to September 2022 (FUY14a) assessment periods, as these time points are when urinary pesticide metabolites, depression and anxiety symptoms were measured. The 2016 FUY8b cohort included 238 participants from the 2008 cohort, and 297 newly recruited adolescent volunteers aged 11-17 years. The 2022 FUY14a follow up included 382 participants from the 2016 FUY8b assessment, and 124 new volunteers. As in 2008, recruitment of new participants was conducted using the System of Local and Community Information, which replaced the SADHS.

Informed consent was obtained from adult participants and child assent was obtained for children 7 years of age or older. ESPINA was approved by the institutional review boards at the University of California San Diego, University of Minnesota, Universidad San Francisco de Quito, UTE University and registered with the Ministry of Public Health of Ecuador.

### Data collection

Participants were examined in local schools for ~2.5 hours during summer closures or on weekends during the school year. Participant height was measured using a height board, and weight was measured using a digital scale (Tanita model 0108MC in 2016; Tanita BF-689 in 2022; Corporation of America, Arlington Heights, IL, USA) (World Health Organization, 2008). Body mass index (BMI) was calculated using the observed height and weight. In FUY-8b BMI-for-age z-score was calculated, and. sexual maturation was assessed using participant self-reports of breast and pubic hair growth for girls, and pubic hair growth for boys using modified drawings of Tanner scales (Emmanuel et al., 2020; Kormorniczak, 2009; Rasmussen et al., 2015). Information on household monthly income, education level, occupation, pesticide use, and other demographic information was collected during interviews with participant parents for FUY8b and with surveys completed by participants for FUY14a (Kornher et al., 2025). Monthly household income was measured continuously for FUY8b, and categorically for FUY 14a. Thus, it was recategorized for both time points into four household income salary groups which were Below Average (less than $500 USD), Average ($500-$999), Above Average ($1000+), and a “Prefer not to answer” group.

### Anxiety Assessments

Anxiety symptoms were measured using age-appropriate scales: the Spanish translated Multidimensional Anxiety Scale for Children 2^nd^ Edition (MASC-2) (Multi-Health Systems [MHS] Inc, North Tonawanda, NY) for the FUY8b and the Spanish version of the Generalized Anxiety Disorder 7 (GAD-7) for the FUY14a. The MASC-2 indexes the range and severity of anxiety symptoms within the last 2 weeks and has been validated in Hispanic children (Fraccaro et al., 2015; Isasi et al., 2014). The MASC-2 English was translated into Spanish with input from community members and was approved by MHS Inc. Completed questionnaires were scored using the MASC-2 Score Software to calculate the scaled T-scores, which are standardized for age and gender (Pearson Canada Assessment, 2020). The T-scores are then classified as Average (<60), Slightly Elevated (60-64), Elevated (65-69), Significantly Elevated (70-74), and Very Elevated (≥75).

The GAD-7 is a seven-item, self-reported questionnaire designed to assess an individual’s anxiety symptoms for the 2 weeks prior to assessment (Spitzer et al., 2006), and has been adapted for Spanish speaking populations. The response scores are classified as Minimal (<=4), Mild (5-9), Moderate (10-14), and Severe (≥15) anxiety symptoms (Pearson Canada Assessment, 2020). This scale has good reliability and validity in English and Spanish (Kroenke et al., 2001; Muñoz-Navarro et al., 2017), and has been administered in Ecuadorian populations (Moreno-Montero et al., 2024; Siteneski et al., 2024).

### Depression Assessments

Depression symptoms were measured using the Children’s Depression Inventory 2^nd^ Edition short assessment (CDI-2), Spanish Version (MHS Inc, North Tonawanda, NY) for the FUY8b and the Spanish Version of the Beck’s Depression Inventory II (BDI-II) for the FUY14a. The CDI-2 short has excellent psychometric properties and yields a score comparable to the full-length version score. The CDI-2 captures symptoms for the 2 weeks prior to the assessment. The CDI-2 Scoring Software scored questionnaires to calculate the scaled T-score, which was standardized for age and gender. The CDI-2 scores are classified as Low (<40), Average (40-59), High Average (60-64), Elevated (65-69) and Very Elevated (70+) depressive symptoms. The BDI-II is a widely used self-report tool for assessing depressive symptom severity within the past 2 weeks (Beck et al., 1996; Dozois et al., 1998; Vázquez Morejón et al., 2014) and has been validated in Spanish-speaking populations(Sánchez-Villena and Farfán Cedrón, 2019), demonstrating reliability across diverse contexts. It consists of 21 items scored on a 4-point Likert scale (0–3), with total scores ranging from 0 to 63. Depression symptom severity is categorized as Minimal (0-13), Mild (14-19), Moderate (20-28), and Severe (29-63).

### Urinary Pesticide Metabolite Measurement

Concentrations of urinary metabolites were measured for a total of 13 insecticides in 2016 and 12 insecticides in 2022 by the U.S. Centers for Disease Control Division of Laboratory Sciences and reported in micrograms per liter (µg/L) (Parajuli et al., 2025). Quality control measures were followed for each step for an optimal robust, reproducible, accurate, precise, and efficient method (Davis et al., 2013). In 2016, to measure organophosphate and pyrethroid metabolite concentrations, targeted metabolites were extracted from one milliliter of urine using a semi-automated solid phase extraction (SPE), removed from each other and other urinary biomolecules with reversed-phase high performance liquid chromatography (HPLC), and quantified with tandem mass spectrometry (MS/MS) via isotope dilution (ID) quantitation (Davis et al., 2013). Neonicotinoid metabolite concentrations were quantified from 0.5 mL of urine using enzymatic hydrolysis of urinary conjugates of target biomarkers, online solid-state extraction, separated by reversed phase HPLC, and detection by ID-electrospray ionization (ES) MS/MS.

For 2022, the organophosphate and pyrethroids metabolites were measured by conducting enzymatic hydrolysis of conjugated metabolites using 0.2 ml of urine, extraction and pre-concentration of the deconjugated metabolites using automated online SPE, and both separation and quantification using HPLC-ID-MS/MS (Wambua et al., 2023). For the neonicotinoids and sulfoximines, we used 0.2 mL of urine, 100 μL of internal standard solution, 300 ul β-glucuronidase solution to hydrolyze conjugated analytes (Baker et al., 2019). The extraction was done with online SPE. A reversed phase Thermo Scientific™ Hypersil Gold aQ column 150 × 4.6 mm, 3 μm particle size, with guard column of the same packing, was used as the analytical column. Quantification was conducted using ID-MS/MS. Strict quality control/quality assurance protocols were followed to ensure data accuracy and reliability for the analytical measurements. All the study samples were re-extracted if quality control failed the statistical evaluation (Baker et al., 2019).

In 2016, analytes included four OP metabolites (para-Nitrophenol [PNP], malathion dicarboxylic acid [MDA], 3,5,6-Trichloro-2-pyridinol [TCPy], & 2-isopropyl-4-methyl-6-hydroxypyrimidine [IMPY]), three PYRs (3-phenoxybenzoic acid [3-PBA], trans-3-(2,2-Dichlorovinyl)-2,2-dimethylcyclopropane carboxylic acid [*trans-*DCCA], & 4-fluoro-3-phenoxybenzoic acid [4F-PBA]) and six neonicotinoid insecticides (5-hydroxy imidacloprid [OHIM], acetamiprid-N-desmethyl [AND], Imidacloprid [IMID], Clothianidin [CLOT], Acetamiprid [ACET], and Thiacloprid [Thiacloprid]). In 2022 PNP, TCPy, IMPY, MDA, 3-PBA, 4PBA, *trans-*DCCA, OHIM, AND, CLOT and two sulfoximines (sulfoxaflor isomer [SLF1] and isomer 2 [SLF2]) were measured. Metabolites were excluded in the analysis if they were detectable in <15%. Thus, CLOT, AND, IMID, 4F-3-PBA, and THIA in 2016, and MDA and 4F-3-PBA in 2022 were excluded.

### Substitution for values below the level of detection (LOD)

For measurements below the LOD, the concentrations were imputed by diving the LOD by the square root of two (Hornung and Reed, 1990).

### Pesticide Summed Variables

Summed exposure values were created for organophosphates, pyrethroids, sulfoximines, neonicotinoids, and overall pesticides. Each summed value was calculated by adding the imputed concentrations of all metabolites measured within that class, regardless detection rates (e.g., 2022 organophosphates summed included PNP, TCPy, IMPY, and MDA). For longitudinal analyses, summed values were restricted to metabolites measured at both time points, ensuring comparability across years.

### Creatinine quantification in urine

Urinary creatinine concentrations were measured by the National Exposure Assessment Laboratory at Emory University to adjust metabolite levels for urinary dilution for the FUY8b, following established methods (Carrieri et al., 2000). A 10-microliter aliquot of each urine sample was diluted and analyzed using HPLC-MS/MS-ES(Kwon et al., 2012). The LOD was 5 mg/dL, and the relative standard deviation (RSD) was 7%.

### Specific gravity quantification in urine

FUY14a specific gravity was measured using an ATAGO PAL-10S digital refractometer (ATAGO Co., Ltd., Tokyo, Japan.) in fresh urine samples at room temperature, following standard procedures. To conduct longitudinal analyses using FUY-8b and FUY-14a, we estimated urinary specific gravity for FUY-8b by converting creatinine values using the Busgang formula that accounted for age and sex designed for children and adolescents. The Busgang formula models the relationship between urinary creatinine and specific gravity via a Gompertz function. Details have been previously published (Busgang et al., 2023).

### Statistical Analysis

#### Descriptive Statistics

Of those assessed, 535 participants in 2016, and 484 participants in 2022 were included after excluding those with missing metabolite measurements, age-appropriate depression and anxiety measures, or covariates (see Linear Models). Between both assessments, there were 646 unique individuals. We calculated the means and standard deviation (STD) for continuous variabels, or count and column percents for categorical variables, stratified by assessment. Geometric means and quantiles were calculated for pesticide metabolite wet weights for both time periods.

#### Linear Models

Generalized Estimating Equations (GEE) assessed associations of pesticide metabolites with depression and anxiety. For metabolites with ≥37% detection, we modeled symptom score differences per interquartile range (IQR) increase in urinary concentration; for metabolites with detection ≥15% but <40%, we also compared participants with detectable (≥LOD) versus non-detectable (<LOD) concentrations. Outcomes were CDI-2 and MASC-2 T-scores at FUY8b and BDI-II and GAD-7 scores at FUY14a. The 2016 model adjusted for age, gender, race, BMI-for-age z-score, tanner score, education, BMI-for-age z-score, and natural log transformed (ln) creatinine. The 2022 models adjusted for age, gender, race, specific gravity, BMI, household monthly income, and education. Covariates were determined a priori.

For longitudinal models, we estimated the anxiety or depression score difference per IQR increase in pesticide metabolite concentrations for metabolites detected in ≥37% of participants in ≥1 follow-up period. If detection was ≥15% but <40% in any follow-up, exposure we also modeled as a binary variable (≥LOD vs. <LOD [reference]). If detection was <15% for any timepoint, it was excluded from the longitudinal models. The longitudinal outcomes were standardized (z-score) anxiety and depression measures. Anxiety outcomes combined MASC-2 and GAD-7 scores, while depression outcomes combined CDI-2 and BDI-II scores. Z-score standardization harmonized results across age-appropriate instruments, which differ in units and variability, enabling consistent linear modeling across follow-ups. MASC-2, GAD-7, CDI-2, and BDI-II z-scores were calculated by subtracting the cohort’s mean score from the participant’s score and dividing it by the standard deviation. Longitudinal models used FUY-14a model covariates. The β (95% confidence intervals [CI]) estimates for the cross-sectional and longitudinal models represented the difference in the anxiety or depression score outcome, for having detectable metabolite concentrations or for an IQR increase in the urinary pesticide metabolite or summary score.

Longitudinal GEE models also estimated whether living with a floricultural or agricultural worker (cohabitation; para-occupational exposure) or working in floriculture or agriculture (agricultural occupation) were associated with the appropriate anxiety and depression outcomes outlined above.

Interaction terms between the pesticide metabolites with gender assessed effect modification by gender. Models were stratified by gender if the interaction term had a p<0.05. A squared pesticide exposure term (e.g., PNP + PNP*PNP) was introduced into the model to assess curvilinearity. For longitudinal analyses, an interaction term between time (FUY8b vs. FUY14a) with pesticide metabolites, agricultural occupation, and cohabitation to assessed time-dependent effects.

#### Logit Models

GEE logit linked models estimated the odds ratio (OR) of having elevated depression (CDI-2≥65 or BDI-2≥20) or anxiety (MASC-2≥60 or GAD-7≥5) symptoms compared to those with less symptoms, for an IQR increase in pesticide metabolite concentrations or summary score (%detectable >37%), or for having detectable levels of the metabolite (15≤%detectable<40%) for FUY8b, FUY14a, and longitudinally (OR 95%CI). Models were also run to assess if cohabitation (ref=did not cohabitate) or agricultural occupation(ref=non-occupational) had greater odds of elevated depression or anxiety symptoms. The covariates were the same as the linear models.

#### Exposure mixture modelling

Quantile g-computation estimated the associations of pesticide biomarker mixtures with depression and anxiety scores cross-sectionally and longitudinally, using the R package “qgcomp”(Keil et al., 2020). Mixture groups for FUY-8b and FUY-14a included metabolites with LOD ≥37%, consisting of (1) an overall pesticide mixture with all such metabolites and (2) separate groups for each pesticide class. In FUY-8b, pyrethroid and neonicotinoid mixtures were not included as only 1 metabolite was detectable in ≥38% in each class. The same was done for the longitudinal models, where mixtures included metabolites that had LOD>37% for ≤1 timepoint. Outcomes were run for linear scores, and for estimation of elevated scores.

Statistically significant associations were defined as p<0.05. Statistical analyses were conducted in SAS 9.4 and R Studio 2024.04.0 for windows.

## Results

### 1. Demographic characteristics

Demographic characteristics are presented in Table 1. The study included 995 observations for 2016 and 2022. Participants were balanced between females (51.4%) and males (48.6%), were predominantly Mestizo or White (80.2%) and had a mean age of 17.3 years (SD = 3.2). The average age was 14.5 (SD=1.8) years in 2016 and 20.3 (SD=1.8) years in 2022. The average BMI was slightly higher in FUY-8b (23.6 [21.6, 25.8]) compared to FUY-14a (20.6 [18.8, 22.4]). Other demographic data did not differ considerably between the two time points. Geometric means and quantiles for pesticide metabolites are presented in Table S1 (FUY-8b), and Table S2 (FUY-14a). Neonicotinoids OHIM, AND, and CLOT detection rates were higher in FUY14a compared to FUY-8b.

**Table 1.** Participant Characteristics, for the July to October 2016 (FUY8b) and July to September 2022 (FUY14a) ESPINA assessments.

|  | Assessment Period |  |
| --- | --- | --- |
|  | 2016 (FUY8b)<br>n=510 | 2022 (FUY14a)<br>n=485 |
| **Age** | 14.46 (1.75) | 20.30 (1.82) |
| **BMI** | 20.91 (2.82) | 24.00 (3.46) |
| **Gender** |  |  |
| Female | 263 (51.6%) | 248 (51.1%) |
| Male | 247 (48.4%) | 237 (48.9%) |
| **Race** |  |  |
| Indigenous | 111 (21.8%) | 86 (17.7%) |
| Mestizo/White | 399 (78.2%) | 399 (82.3%) |
| **Lives with a Floricultural or Agricultural Worker** |  |  |
| No, n (%) | 166 (32.6%) | 131 (27.0%) |
| Yes, n (%) | 344 (67.5%) | 354 (73.0%) |
| **Works in Floriculture or Agriculture** |  |  |
| No, n (%) | 479 (93.9%) | 352 (72.6%) |
| Yes, n (%) | 31 (6.1%) | 133 (27.4%) |
| **Monthly Household Income (USD)** |  |  |
| Below Average: Less than \$500, n (%) | 222 (43.5%) | 253 (52.2%) |
| Average: \$500 to \$999, n (%) | 247 (48.4%) | 104 (21.4%) |
| Above Average: \$1000+, n (%) | 41 (8.0%) | 33 (6.8%) |
| Prefer Not to Answer, n (%) | 0 (0.0%) | 95 (19.6%) |
| **Education** |  |  |
| Elementary and Middle School, n (%) | 146 (28.6%) | 10 (2.1%) |
| Some High School, n (%) | 364 (71.4%) | 63 (13.0%) |
| High School Grad, n (%) | 0 (0.0%) | 220 (45.4%) |
| Some College and Above, n (%) | 0 (0.0%) | 192 (39.6%) |
| **Depression Scales** |  |  |
| Depression z-score | 0.00 (1.00) | 0.00 (1.00) |
| Prevalence of elevated scores:<br>(CDI>65 or BDI2>=20), n (%) | 47 (9.36%) | 83 (17.11%) |
| CDI-2 T-score | 53.09 (9.38) |  |
| CDI-2 Categories |  |  |
| Low (<40), n (%) | 18 (3.59%) |  |
| Average (40-59), n (%) | 370 (73.71%) |  |
| High Average(60-64), n (%) | 67 (13.35%) |  |
| Elevated (65-69), n (%) | 21 (4.18%) |  |
| Very Elevated (70+), n (%) | 26 (5.18%) |  |
| BDI-II raw score |  | 10.38 (9.61) |
| BDI-II Categories |  |  |
| Minimal (0-13), n (%) |  | 330 (68.04%) |
| Mild (14-19), n (%) |  | 72 (14.85%) |
| Moderate (20-28), n (%) |  | 53 (10.93%) |
| Severe (29-63), n (%) |  | 30 (6.19%) |
| **Anxiety Scales** |  |  |
| Anxiety z-score | 0.00 (1.00) | 0.00 (1.00) |
| Prevalence of elevated scores (MASC≥60 or GAD7≥5), n (%) | 223 (43.73%) | 216 (44.54%) |
| MASC-II T-Score | 57.64 (9.57) |  |
| MASC-II Categories |  |  |
| Average (<55), n (%) | 185 (36.63%) |  |
| Slightly Elevated (55-59), n (%) | 97 (19.21%) |  |
| Elevated (60-64), n (%) | 108 (21.39%) |  |
| Significantly Elevated (65-69), n (%) | 52 (10.30%) |  |
| Very Elevated (≥70), n (%) | 63 (12.48%) | |
| GAD-7 Total Score |  | 4.86 (4.20) |
| GAD-7 Full Categories |  |  |
| Minimal (≥4), n (%) | | 269 (55.46%) |
| Mild Anxiety (5-9), n (%) |  | 154 (31.75%) |
| Moderate Anxiety (10-14), n (%) |  | 43 (8.87%) |
| Severe Anxiety (15+), n (%) |  | 19 (3.92%) |

BDI-II = Becks Depression Inventory-II, CDI-2 = Children's Depression Inventory-2, GAD-7 = General Anxiety Disorder-7, MASC-2 = Multidimensional Anxiety Scale for Children-2 Values presented are mean (SD) or N (percent)

### 2. Longitudinal analyses (FUY-8b, FUY-14a) of urinary metabolites with anxiety and depression scores

#### Linear models

Accounting for time, most urinary pesticide summed scores, but not the metabolites, were consistently associated with depression (Table 2). Overall pesticides (β=0.06 [0.01, 0.11]), organophosphates (β=0.05 [0.001, 0.10]), and pyrethroid (β=0.03 [0.01, 0.04]) summed concentrations were positively associated with depression. For the metabolites, an IQR higher TCPy concentration was associated with 0.06 higher depression symptoms (β=0.06, 95% CI[0.02, 0.10]). A small positive association was also observed for *trans*-DCCA with depression (β=0.01, [ 0.003, 0.02]). No statistically significant associations were found for anxiety. Time interactions were observed for associations between the depression z-score with overall pesticides summed, organophosphates summed, TCPy and OHIM, and between anxiety z-score with AND, which prompted further cross-sectional analyses. No gender interactions or curvilinearity were observed.

**Table 2.**
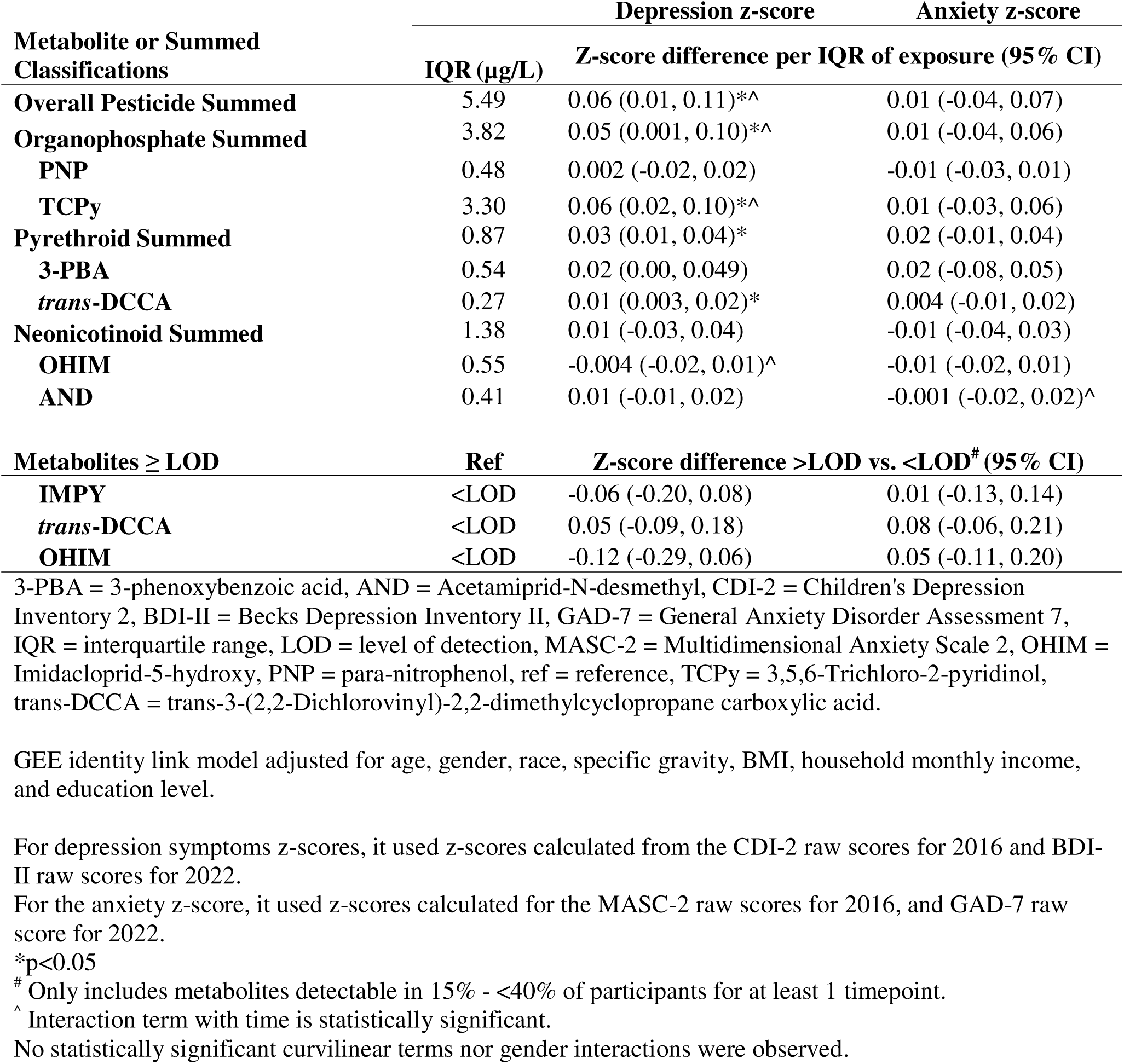
Longitudinal associations in FUY-8b (2016) and FUY-14a (2022) of urinary insecticide metabolites with depression and anxiety z-scores in adolescents and young adults of the ESPINA study (n=646, observations=995).

#### Logit models

For the longitudinal logit link GEE model (Figure 1), no statistically significant associations were observed between the classification summed concentrations and depression. Among individual metabolites, only higher TCPy concentrations were associated with greater odds of elevated depression (OR=1.14, 95% CI:[1.02, 1.28]). For anxiety, most exposures were not associated with elevated symptoms. However, detectable trans-DCCA (OR=1.39, [1.03, 1.87]) and OHIM (OR=1.44, [1.02, 2.04]) were linked to higher odds of elevated anxiety.

**Figure 1.**
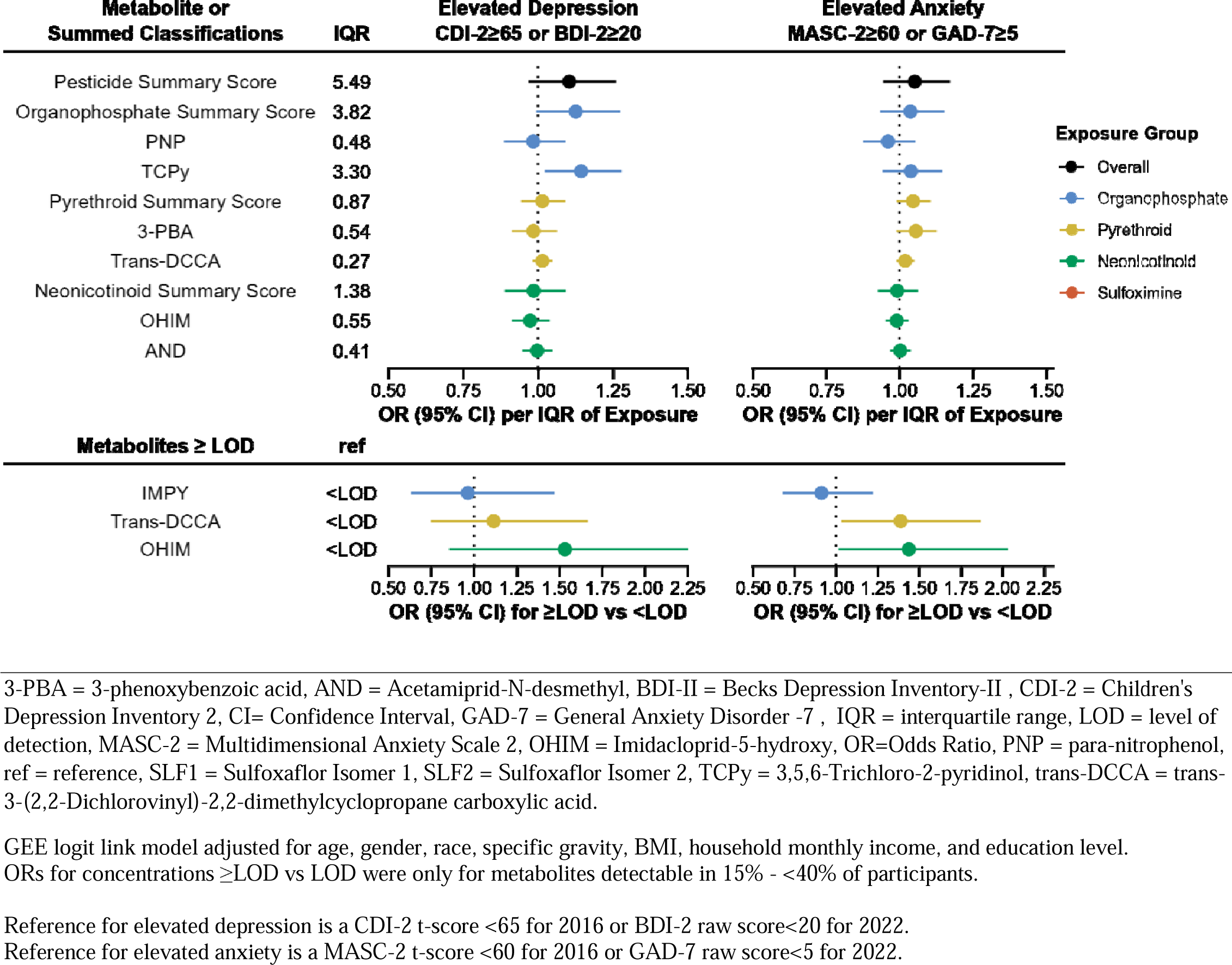
Forest plot of longitudinal associations in FUY-8b (2016) and FUY-14a (2022) of urinary insecticide metabolites with odds of elevated depression and anxiety scores among adolescents and young adults in the ESPINA study (n=646, observations=995).

### 3. Longitudinal analyses (FUY-8b, FUY-14a) of occupational and para-occupational exposures with anxiety and depression scores

For depression outcomes, neither cohabitation (β=0.01, 95% CI: –0.13, 0.15) nor agricultural occupation (β=0.05, 95% CI: [–0.13, 0.23]) was associated with depression z-scores. Similarly, in logistic models, neither cohabitation (OR=1.07 [0.7, 1.64]) nor agricultural occupation (OR=1.07 [0.64, 1.79]) was associated with elevated depression symptoms.

For anxiety outcomes, cohabitation (β=0.02, [–0.12, 0.15]) and agricultural occupation (β=0.03 [–0.14, 0.20) were not associated with anxiety z-scores. Logistic models also showed no associations with elevated anxiety symptoms for cohabitation (OR=0.97 [0.73, 1.30]) nor agricultural occupation (OR=1.13 [0.77, 1.64]). There were no interactions with time, thus no cross-sectional models were run for cohabitation and occupation.

### 4. Cross-sectional analyses (FUY-8b, 2016) of urinary metabolites with anxiety and depression scores

Table 3 presents linear associations for FUY-8b. We observed significant positive associations between overall pesticides summed (β=1.00 [1.01 [0.48, 1.54]), organophosphates summed (β=0.99 [0.47, 1.51]), TCPy (β=0.97 95% CI:[0.49, 1.45]), the pyrethroids summed (β=0.25 [0.10, 0.40]) and 3-PBA (β= 0.31 [ 0.01, 0.61]) with CDI-2 t-scores. Compared to individuals with *trans*-DCCA<LOD, individuals with *trans*-DCCA concentration ≥LOD had 3.34 (95% CI: 1.23, 5.45) higher MASC-2 T-scores. Having detectable MDA lowered depression scores (β=−1.69 [−3.33, −0.04]). No statistically significant gender interactions were observed; thus, the models were not stratified.

**Table 3.**
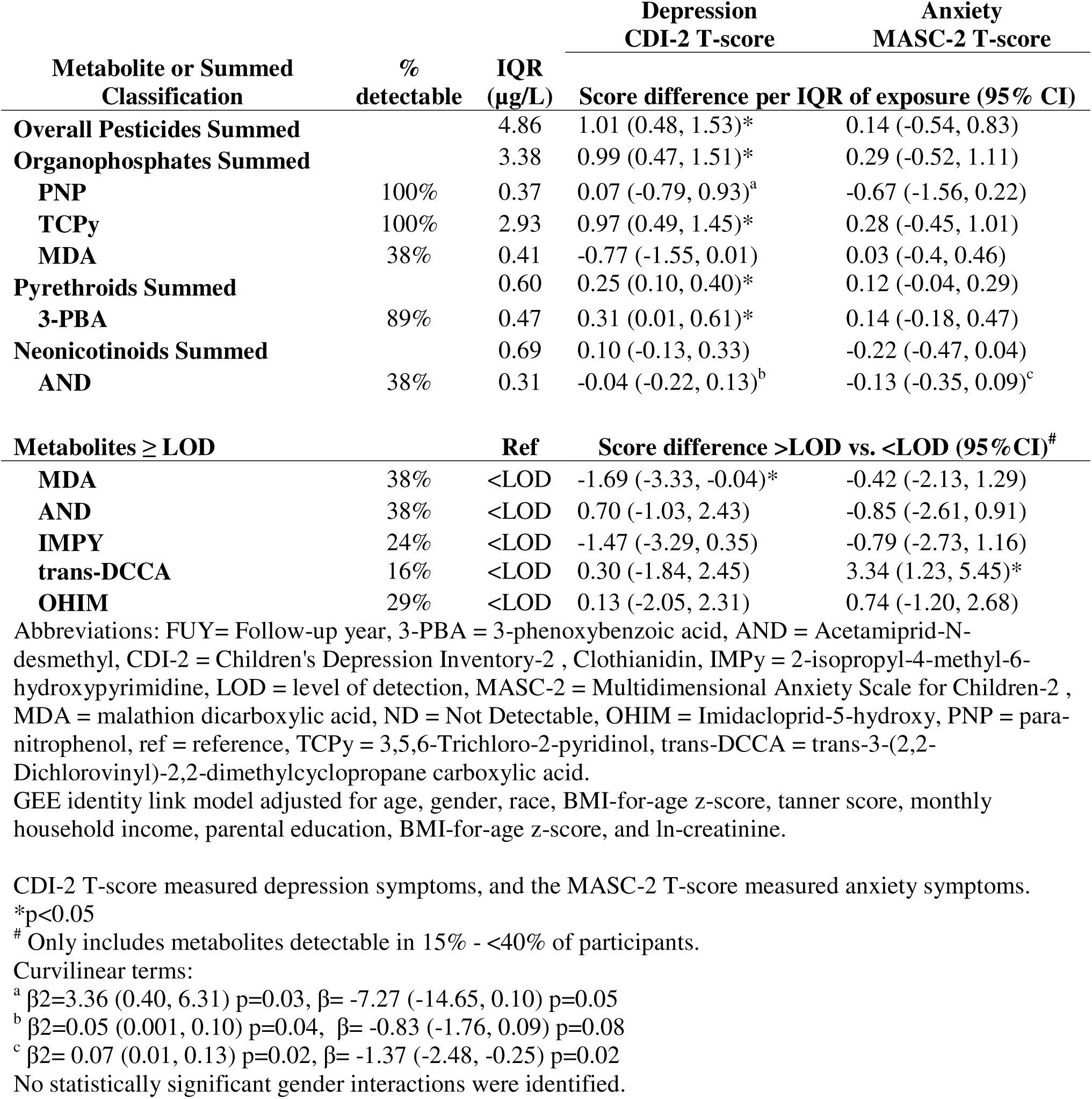
Cross-sectional associations in FUY-8b (2016) of urinary insecticide metabolites with depression and anxiety scores in adolescents of the ESPINA study (n=510).

Figure 2 presents a forest plot of the FUY-8b GEE logit models. Detectable OHIM concentrations were linked to two-fold higher odds of elevated depression symptoms (OR = 1.99, 95% CI: 1.01, 3.94). For anxiety, detectable trans-DCCA concentrations had 152% greater odds of elevated anxiety (OR = 2.52, 95% CI: 1.52, 4.16).

**Figure 2.**
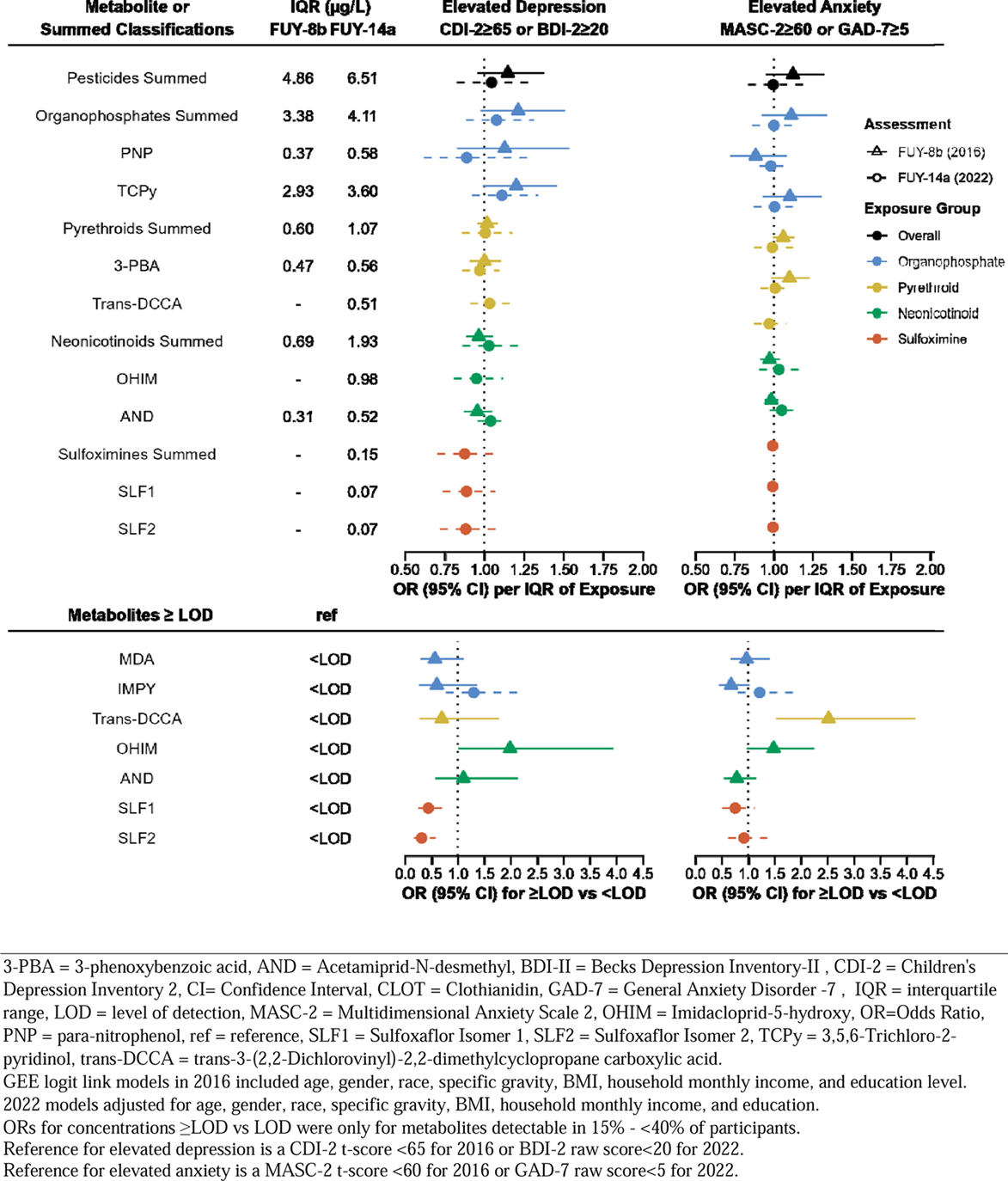
Forest plot of cross-sectional associations in FUY-8b (2016; n=510) and FUY-14a (2022; n=485) of urinary insecticide metabolites with depression and anxiety scores in adolescents and adults of the ESPINA study.

### 5. Cross-sectional analyses (FUY-14a, 2022) of urinary metabolites with anxiety and depression scores

For FUY-14a GEE identity link models (Table 4), negative associations with BDI-II total scores were noted for the sulfoximines summed (β=−1.42 [−2.14, −0.69]), SLF1 (β=−0.11 [−0.18, −0.03]) and SLF2 (β=−0.12 [−0.20, −0.04]). Having SLF1 concentrations >LOD lowered odds of elevated GAD-7 (β=−0.81 [−1.56, −0.07]). No gender interactions or curvilinearity was observed.

**Table 4.** Cross-sectional associations in FUY-14a (2022) of urinary insecticide metabolites with depression and anxiety scores in adolescents and young adults of the ESPINA study (n=485).

| Metabolite or Summed Classification | % detectable | IQR (µg/L) | Depression<br>BDI-II Total Score | Anxiety<br>GAD-7 Total Score |
| --- | --- | --- | --- | --- |
|  |  |  | Score difference per IQR of exposure (95%CI) |  |
| **Overall Pesticide Summed** |  | 6.51 | 0.03 (-0.8, 0.86) | 0.04 (-0.35, 0.42) |
| **Organophosphates Summed** |  | 4.11 | 0.19 (-0.55, 0.92) | 0.05 (-0.25, 0.35) |
| **PNP** | 100% | 0.58 | 0.01 (-0.17, 0.19) | -0.02 (-0.1, 0.06) |
| **TCPy** | 100% | 3.60 | 0.30 (-0.41, 1.01) | 0.04 (-0.24, 0.32) |
| **Pyrethroids Summed** |  | 1.07 | 0.11 (-0.44, 0.66) | 0.06 (-0.22, 0.34) |
| **3-PBA** | 99% | 0.56 | 0.14 (-0.26, 0.55) | 0.11 (-0.10, 0.32) |
| **trans-DCCA** | 43% | 0.51 | -0.02 (-0.48, 0.44) | -0.05 (-0.28, 0.18) |
| **Neonicotinoids Summed** |  | 1.93 | 0.06 (-0.51, 0.63) | 0.08 (-0.22, 0.38) |
| **OHIM** | 93% | 0.98 | -0.14 (-0.48, 0.20) | -0.03 (-0.22, 0.15) |
| **AND** | 95% | 0.52 | 0.24 (-0.07, 0.55) | 0.14 (-0.01, 0.29) |
| **CLOT** | 74% | 0.77 | 0.58 (-0.33, 1.48) | 0.35 (-0.12, 0.82) |
| **Sulfoximines Summed** |  | 0.15 | -0.12 (-0.2, -0.04)** | -0.03 (-0.07, 0.003) |
| **SLF1** | 38% | 0.07 | -0.11 (-0.18, -0.03)* | -0.03 (-0.06, 0.01) |
| **SLF2** | 37% | 0.07 | -0.12 (-0.20, -0.04)* | -0.03 (-0.06, 0.01) |
| **Metabolites ≥ LOD** |  | **Ref** | **Score difference >LOD vs. <LOD (95% CI)<sup>#</sup>** |  |
| **IMPY** | 29% | <LOD | 0.23 (-1.61, 2.08) | 0.41 (-0.41, 1.24) |
| **SLF1** | 38% | <LOD | -2.20 (-3.93, -0.47)* | -0.81 (-1.56, -0.07)* |
| **SLF2** | 37% | <LOD | -2.77 (-4.42, -1.12)* | -0.48 (-1.21, 0.25) |

3-PBA = 3-phenoxybenzoic acid, AND = Acetamiprid-N-desmethyl, BDI-II = Becks Depression Inventory-II, CLOT = Clothianidin, GAD-7 = General Anxiety Disorder -7, IMID = Imidacloprid-5-hydroxy, IQR = interquartile range, LOD = level of detection, OHIM = Imidacloprid-5-hydroxy, PNP = para-nitrophenol, ref = reference, SLF1 = Sulfoxaflor Isomer 1, SLF2 = Sulfoxaflor Isomer 2, TCPy = 3,5,6-Trichloro-2-pyridinol, trans-DCCA = trans-3-(2,2-Dichlorovinyl)-2,2-dimethylcyclopropane carboxylic acid.
GEE identity link models adjusted for age, gender, race, specific gravity, BMI, household monthly income, and education.
Depression symptoms were measured using the BDI-II total scores, and anxiety symptoms were measured using the GAD-7 total scores.
\*p<0.05
<sup>#</sup> Only includes metabolites detectable in 15% - <40% of participants.
No statistically significant curvilinear nor gender interaction terms were identified.

For the GEE logit models (Figure 2), an IQR higher clothianidin concentration was associated with greater odds of elevated depression (OR=1.41, [1.08, 1.84]) and anxiety symptoms (OR=1.44 [1.07, 1.93]). In contrast, having detectable SLF1 (0.44 [0.25, 0.78]) and SLF2 (OR=0.32 [0.17, 0.58]) concentrations lowered the odds of elevated depression.

### 6. Pesticide mixtures in relation to depression and anxiety scores

Quantile g-computation analyses indicated few consistent associations between pesticide mixtures and depression or anxiety outcomes across time points (Supplemental Table 3). In the 2016 cross-sectional assessment, no significant associations were observed for overall or class-specific mixtures with depression (CDI-2) or anxiety (MASC-2). In the 2022 cross-sectional assessment, a quartile increase in sulfoxaflor mixture exposure was inversely associated with both depression (BDI-II: Ψ = –3.01, 95% CI: –5.01, –1.01) and anxiety (GAD-7: Ψ = –0.99, 95% CI: –1.86, –0.14). Logistic models further showed that sulfoxaflor mixture was linked to lower odds of elevated depression in 2022 (OR = 0.54, 95% CI: 0.38, 0.77), while neonicotinoid mixtures were associated with greater odds of elevated anxiety (OR = 1.21, 95% CI: 1.05, 1.40). In longitudinal analyses which excluded SLF1 and SLF2, no significant associations were identified in any of the analyses. Weights values varied depending on the mixture (Figure S1).

## Discussion

This study of Ecuadorian adolescents and young adults suggests that exposure to organophosphate, pyrethroid, and neonicotinoid pesticides is associated with higher depression and anxiety symptoms, particularly among adolescents. In 2016, overall pesticides summed, organophosphate summed, TCPy, pyrethroids summed, 3-PBA, and OHIM were linked to greater depression symptoms, while trans-DCCA was associated with anxiety. In 2022, sulfoximine metabolites were inversely associated with depression, whereas CLOT was positively associated with both depression and anxiety. Longitudinal analyses confirmed that higher overall pesticides summed, organophosphates summed, TCPy, pyrethroids summed and trans-DCCA were related to greater depression symptoms. No internalizing symptom score differences were observed with work in agriculture, cohabitation with agricultural workers nor gender. This is among the first studies to report longitudinal links between urinary pesticide metabolites and internalizing symptoms from adolescence into adulthood.

The associations observed between pyrethroid, organophosphate and neonicotinoid metabolites with higher depression symptoms are consistent with published literature. Pesticide exposure has been linked to higher depression levels in occupational and non-occupational adults in the United States, Brazil, and France (Beseler and Stallones, 2008; Conti et al., 2018; Darney et al., 2018). In Korean adults, a higher proportion of individuals reporting depression also reported using pesticides (Koh et al., 2017). Geriatric adults living near known pyrethroid application locations had greater odds of depression and anti-depressant use (Furlong et al., 2020). Prenatal pyrethroid exposure, measured by 3-PBA, has also been linked to worse childhood internalizing scores (Furlong et al., 2017). The same study found detectable prenatal *trans-*DCCA was positively associated with anxiety but not depression, consistent with our 2016 findings.

For organophosphates, the association between organophosphates summed and TCPy with depression is supported by prior research. Rodent models demonstrated repeated chlorpyrifos (TCPy parent chemical) exposure altered behavioral performance indicative of worse emotional wellbeing (Chen et al., 2011). In the 2016 ESPINA cohort, lower AChE activity, indicative of higher organophosphate exposure, was associated with higher depression symptoms, strongest in girls and younger children (Suarez-Lopez et al., 2020, 2019). Human studies have also reported mood changes following organophosphate pesticide poisonings (Khan et al., 2019; London et al., 2005). No associations were observed for PNP, and detectable MDA was negatively associated with depression in 2016, despite positive associations in the literature. Clinical reports have linked high level parathion exposure (PNP parent chemical) to increased depressive and anxiety symptoms in clinical cases (Clifford and Nies, 1989), and findings from rodent models observed that repeated exposure to malathion induced depressive-like behavior in rodents (Saeedi Saravi et al., 2016). Parathion and malathion exposure in ESPINA may be under thresholds needed to affect mood, particularly since parathion has been banned in Ecuador since 2009 and ethyl, diethyl, and methyl parathion since 1992 (Agencia de Regulación y Control Fito y Zoosanitario, 2025).

Animal studies also suggest neonicotinoids affect mood. Imidacloprid (OHIM parent chemical) altered *drosophila melanogaster* behaviors (Janner et al., 2021). CLOT increased anxiety-like behaviors even at doses below the no observed adverse effect level (Hirano et al., 2018). The positive association between detectable OHIM with elevated depression score in 2016, and CLOT concentration with elevated anxiety and depression in 2022 support these findings. This is the first study to assess and find neonicotinoid mixture effects on elevated anxiety symptoms in an adolescent and adult population, warranting future research.

The consistent negative association between sulfoximines (SLF1 and SLF2) and depression symptoms in 2022 may reflect their strong effects on nAChRs. In vitro, sulfoxaflor produced large-amplitude nAChR currents, greater than those elicited by several neonicotinoids but not clothianidin (Watson et al., 2011)These receptor-level effects suggest sulfoximines, like nicotine, may influence mood-regulating pathways. nAChRs are activated when acetylcholine or nicotine binds between α–α or α–β subunits, opening the channel to sodium and calcium influx (Galzi and Changeux, 1995; Miyazawa et al., 2003). Nicotine consumption has been shown to alleviate depression symptoms (Picciotto et al., 2015), and antidepressants also act as nAChR antagonists, with SSRIs and NRIs demonstrating greater efficacy when combined with nAChR antagonists or partial agonists (Andreasen et al., 2009). Because sulfoxaflor has stronger nAChR affinity than many neonicotinoids, this may explain why SLF1 and SLF2 were linked to fewer depression symptoms, while neonicotinoids were not.

Some null associations may reflect chronic low-level exposure among many ESPINA participants insufficient to elicit mood-related effects. Participants were generally not pesticide applicators (6.08% in 2016, and 27.42% in 2022), and exposures were generally lower than those seen in poisoning cases. Nonetheless, exposure levels in this population are considered higher than those of urban populations (Mehta et al., 2025).

In general, we observed more pronounced associations between organophosphate and pyrethroid insecticide metabolites with depression and anxiety symptoms in adolescence than during young adulthood. Compared to adults, adolescents exhibit increased sensitivity to pesticide-related toxicants, which elevates their risk for adverse health outcomes with potential long-term consequences(Barker, 2004, p. 2; Landrigan and Goldman, 2011; Roberts et al., 2012; Suk et al., 2016). Thus, adolescence may be a sensitive age where pesticide exposure more strongly impacts mental health. To our knowledge, no prior studies have examined the effects of pesticide exposure on mental health from adolescence into adulthood, and additional research is needed to determine whether our findings are consistent across other populations.

Strengths of this analysis include the large adolescent and young adult cohort assessed across two collection periods spanning six years, using standardized internalizing symptom scales, and using measured pesticide metabolites to estimate pesticide exposure rather than relying on self-reported exposure. This study expands mental health research of Latin American populations, which is under-represented in the scientific literature. Limitations include the inconsistent, but age-appropriate, scales used at both time periods but we used a z-score to allow longitudinal modeling. A limitation for neonicotinoids is that the analytical methods used in 2016 had nearly twice the detection limits compared to 2022, resulting in lower detectability. This reduced the number of quantifiable associations and decreased confidence in the estimates that could be measured. Medication use may have biased findings. Medication use was not collected in 2016, and only 1 individual reported using a serotonin-norepinephrine reuptake inhibitor in 2022. Under-reporting of anxiolytic or antidepressant use may bias results toward the null, as these medications can reduce depression and anxiety symptoms, thereby diminishing the apparent associations between pesticide exposures and internalizing outcomes.

## Conclusion

Depression and anxiety are the most prevalent psychiatric disorders in the world, underscoring the importance of identifying potential risk factors (Kessler et al., 2005; Sansone and Sansone, 2010; World Health Organization, 2017). This paper identified positive associations between organophosphates (TCPy), pyrethroids (3-PBA, trans-DCCA), and neonicotinoids (OHIM, CLOT) with depression and anxiety symptoms, in adolescence and young adulthood. Longitudinal analyses further sustained associations of TCPy, trans-DCCA, and OHIM with internalizing symptoms from during the transition from adolescence to young adulthood of a Latin American population. As more positive associations between pesticide metabolites with depression and anxiety were observed in the adolescent cross-section, it suggests greater sensitivity to pesticide exposure on mental health at this age. Sulfoximine metabolites were protective against anxiety and depression, potentially due to their affinity to nAChRs mood regulating pathways. Because pesticides use is pervasive across the world, these findings highlight the need for further guidance to develop policies that would limit pesticide exposure and consequently protect agricultural populations, especially early in the developmental period.

## Data Availability

All data produced in the present study are available upon reasonable request to the authors.

## Acknowledgements

We thank Fundación Cimas del Ecuador, the Parish Governments of Pedro Moncayo County, community members of Pedro Moncayo and the Education District of Pichincha-Cayambe-Pedro Moncayo counties for their support on this project. We thank Dr. Maria Ospina and Dr. Antonia M. Calafat for their contributions in quantifying most urinary pesticide metabolites. We thank Naimisha Adira on her editorial assistance, and Dr. Carlos Gould for his valuable guidance on the development and refinement of the figures included in this manuscript.

## Declaration of generative AI and AI-assisted technologies in the manuscript preparation process

During the preparation of this work the authors used ChatGPT for analytic code troubleshooting and manuscript editing. After using this tool, the authors reviewed and edited the content as needed and take full responsibility for the content of the published article.

## Ethical Considerations

Informed consent of adult participants was obtained. For minor participants (<18 years of age), we obtained parental or legal guardian’s authorization for participation of their children and child assent was obtained for all children who were 7 years of age and older. This study was approved by the institutional review boards at the University of California San Diego, University of Minnesota, Universidad San Francisco de Quito, and UTE University. This study was also registered at the Ministry of Public Health of Ecuador.

## References

Agencia de Regulación y Control Fito y Zoosanitario, 2025. LISTADO DE PLAGUICIDAS PROHIBIDOS EN EL ECUADOR. Agencia de Regulación y Control Fito y Zoosanitario.

Andreasen, J.T., Olsen, G.M., Wiborg, O., Redrobe, J.P., 2009. Antidepressant-like effects of nicotinic acetylcholine receptor antagonists, but not agonists, in the mouse forced swim and mouse tail suspension tests. Journal of Psychopharmacology 23, 797–804. 10.1177/0269881108091587

Baker, S.E., Serafim, A.B., Morales-Agudelo, P., Vidal, M., Calafat, A.M., Ospina, M., 2019. Quantification of DEET and neonicotinoid pesticide biomarkers in human urine by online solid-phase extraction high-performance liquid chromatography-tandem mass spectrometry. Analytical and Bioanalytical Chemistry 411, 669–678. 10.1007/s00216-018-1481-0

Barker, D.J.P., 2004. The developmental origins of adult disease. J Am Coll Nutr 23, 588S–595S. 10.1080/07315724.2004.10719428

Beck, A.T., Steer, R.A., Brown, G.K., 1996. Beck depression inventory.

Beseler, C.L., Stallones, L., 2008. A Cohort Study of Pesticide Poisoning and Depression in Colorado Farm Residents. Annals of Epidemiology 18, 768–774. 10.1016/j.annepidem.2008.05.004

Beseler, C.L., Stallones, L., Hoppin, J.A., Alavanja, M.C.R., Blair, A., Keefe, T., Kamel, F., 2008. Depression and pesticide exposures among private pesticide applicators enrolled in the Agricultural Health Study. Environmental health perspectives 116, 1713–9. 10.1289/ehp.11091

Beseler, C.L., Stallones, L., Hoppin, J.A., Alavanja, M.C.R., Blair, A., Keefe, T., Kamel, F., 2006. Depression and pesticide exposures in female spouses of licensed pesticide applicators in the agricultural health study cohort. Journal of occupational and environmental medicine / American College of Occupational and Environmental Medicine 48, 1005–13. 10.1097/01.jom.0000235938.70212.dd

Busgang, S.A., Andra, S.S., Curtin, P., Colicino, E., Mazzella, M.J., Bixby, M., Sanders, A.P., Meeker, J.D., Hauptman, M., Yelamanchili, S., Phipatanakul, W., Gennings, C., 2023. A cross-validation based approach for estimating specific gravity in elementary-school aged children using a nonlinear model. Environmental Research 217. 10.1016/j.envres.2022.114793

Carrieri, M., Trevisan, A., Bartolucci, G.B., 2000. Adjustment to concentration-dilution of spot urine samples: Correlation between specific gravity and creatinine. International Archives of Occupational and Environmental Health 74, 63–67. 10.1007/s004200000190

Chen, W.Q., Yuan, L., Xue, R., Li, Y.F., Su, R.B., Zhang, Y.Z., Li, J., 2011. Repeated exposure to chlorpyrifos alters the performance of adolescent male rats in animal models of depression and anxiety. NeuroToxicology 32, 355–361. 10.1016/j.neuro.2011.03.008

Clifford, N.J., Nies, A.S., 1989. Organophosphate Poisoning From Wearing a Laundered Uniform Previously Contaminated With Parathion. JAMA: The Journal of the American Medical Association 262, 3035–3036. 10.1001/jama.1989.03430210077034

Conti, C.L., Barbosa, W.M., Simão, J.B.P., Álvares-da-Silva, A.M., 2018. Pesticide exposure, tobacco use, poor self-perceived health and presence of chronic disease are determinants of depressive symptoms among coffee growers from Southeast Brazil. Psychiatry Research 260, 187–192. 10.1016/j.psychres.2017.11.063

Darney, K., Bodin, L., Bouchard, M., Côté, J., Volatier, J.L., Desvignes, V., 2018. Aggregate exposure of the adult French population to pyrethroids. Toxicology and Applied Pharmacology 351, 21–31. 10.1016/j.taap.2018.05.007

Davis, M.D., Wade, E.L., Restrepo, P.R., Roman-Esteva, W., Bravo, R., Kuklenyik, P., Calafat, A.M., 2013. Semi-automated solid phase extraction method for the mass spectrometric quantification of 12 specific metabolites of organophosphorus pesticides, synthetic pyrethroids, and select herbicides in human urine. Journal of Chromatography B: Analytical Technologies in the Biomedical and Life Sciences 929, 18–26. 10.1016/j.jchromb.2013.04.005

Dozois, D.J.A., Dobson, K.S., Ahnberg, J.L., 1998. A psychometric evaluation of the Beck Depression Inventory–II. Psychological Assessment 10, 83–89. 10.1037/1040-3590.10.2.83

Ecobichon, D.J., 2001. Pesticide use in developing countries. Toxicology 160, 27–33. 10.1016/S0300-483X(00)00452-2

Emmanuel, M., Bokor, B.R., Bornstein, M.H., 2020. Tanner Stages, The SAGE Encyclopedia of Lifespan Human Development. StatPearls Publishing, Treasure Island (FL). 10.4135/9781506307633.n814

Ferré, D.M., Quero, A.A.M.M., Hernández, A.F., Hynes, V., Tornello, M.J., Lüders, C., Gorla, N.B.M.M., Ferre, D.M., Quero, A.A.M.M., Hernandez, A.F., Hynes, V., Tornello, M.J., Luders, C., Gorla, N.B.M.M., 2018. Potential risks of dietary exposure to chlorpyrifos and cypermethrin from their use in fruit/vegetable crops and beef cattle productions. Environmental monitoring and assessment 190, 292. 10.1007/s10661-018-6647-x

Fraccaro, R.L., Stelnicki, A.M., Nordstokke, D.W., 2015. Test Review: Multidimensional Anxiety Scale for Children by J. S. March. Canadian Journal of School Psychology 30, 70–77. 10.1177/0829573514542924

Furlong, M.A., Barr, D.B., Wolff, M.S., Engel, S.M., 2017. Prenatal exposure to pyrethroid pesticides and childhood behavior and executive functioning. NeuroToxicology 62, 231–238. 10.1016/j.neuro.2017.08.005

Furlong, M.A., Paul, K.C., Cockburn, M., Bronstein, J., Keener, A., Rosario, I.D., Folle, A.D., Ritz, B., 2020. Ambient Pyrethroid Pesticide Exposures in Adult Life and Depression in Older Residents of California’s Central Valley. Environmental Epidemiology 4, e123–e123. 10.1097/ee9.0000000000000123

Galzi, J.-L., Changeux, J.-P., 1995. Neuronal Nicotinic Receptors: Molecular Organization and Regulations (No. 0028-3908(95)00034–8), Neuropharmacology.

Harrison, V., Mackenzie Ross, S., 2016. Anxiety and depression following cumulative low-level exposure to organophosphate pesticides. Environmental Research 151, 528–536. 10.1016/j.envres.2016.08.020

Hirano, T., Yanai, S., Takada, T., Yoneda, N., Omotehara, T., Kubota, N., Minami, K., Yamamoto, A., Mantani, Y., Yokoyama, T., Kitagawa, H., Hoshi, N., 2018. NOAEL-dose of a neonicotinoid pesticide, clothianidin, acutely induce anxiety-related behavior with human-audible vocalizations in male mice in a novel environment. Toxicology Letters 282, 57–63. 10.1016/j.toxlet.2017.10.010

Hornung, R.W., Reed, L.D., 1990. Estimation of Average Concentration in the Presence of Nondetectable Values Estimation of Average Concentration in the Presence of Nondetectable Values. Appl. Occup. Environ. Hyg 5, 46–51. 10.1080/1047322X.1990.10389587

Isasi, C.R., Carnethon, M.R., Ayala, G.X., Arredondo, E., Bangdiwala, S.I., Daviglus, M.L., Delamater, A.M., Eckfeldt, J.H., Perreira, K., Himes, J.H., Kaplan, R.C., Van Horn, L., 2014. The Hispanic Community Children’s Health Study/Study of Latino Youth (SOL Youth): Design, objectives, and procedures. Annals of Epidemiology 24, 29–35. 10.1016/j.annepidem.2013.08.008

Janner, D.E., Gomes, N.S., Poetini, M.R., Poleto, K.H., Musachio, E.A.S., de Almeida, F.P., de Matos Amador, E.C., Reginaldo, J.C., Ramborger, B.P., Roehrs, R., Prigol, M., Guerra, G.P., 2021. Oxidative stress and decreased dopamine levels induced by imidacloprid exposure cause behavioral changes in a neurodevelopmental disorder model in Drosophila melanogaster. NeuroToxicology 85, 79–89. 10.1016/j.neuro.2021.05.006

Keil, A.P., Buckley, J.P., O’Brien, K.M., Ferguson, K.K., Zhao, S., White, A.J., 2020. A Quantile-Based g-Computation Approach to Addressing the Effects of Exposure Mixtures. Environmental Health Perspectives. 10.1289/EHP5838

Kessler, R.C., Berglund, P., Demler, O., Jin, R., Merikangas, K.R., Walters, E.E., 2005. Lifetime prevalence and age-of-onset distributions of DSM-IV disorders in the national comorbidity survey replication. Archives of General Psychiatry 62, 593–602. 10.1001/archpsyc.62.6.593

Khan, N., Kennedy, A., Cotton, J., Brumby, S., 2019. A pest to mental health? Exploring the link between exposure to agrichemicals in farmers and mental health. International Journal of Environmental Research and Public Health 16. 10.3390/ijerph16081327

King, A.M., Aaron, C.K., 2015. Organophosphate and Carbamate Poisoning. Emergency Medicine Clinics of North America 33, 133–151. 10.1016/j.emc.2014.09.010

Koh, S.B., Kim, T.H., Min, S., Lee, K., Kang, D.R., Choi, J.R., 2017. Exposure to pesticide as a risk factor for depression: A population-based longitudinal study in Korea. NeuroToxicology 62, 181–185. 10.1016/j.neuro.2017.07.005

Kormorniczak, M., 2009. Tanner Scale. Wikipedia. 10.32388/8rd4dl

Kornher, K., Gould, C.F., Manzano, J.M., Baines, K., Kayser, G., Tu, X., Suarez-Torres, J., Martinez, D., Peterson, L.A., Huset, C.A., Barr, D.B., Suarez-Lopez, J.R., 2025. Associations of PFAS and pesticides with lung function changes from adolescence to young adulthood in the ESPINA study. International Journal of Hygiene and Environmental Health 265, 114526. 10.1016/j.ijheh.2025.114526

Kroenke, K., Spitzer, R.L., Williams, J.B.W., 2001. The PHQ-9: Validity of a brief depression severity measure. Journal of General Internal Medicine 16, 606–613. 10.1046/j.1525-1497.2001.016009606.x

Kwon, W., Kim, J.Y., Suh, S., In, M.K., 2012. Simultaneous determination of creatinine and uric acid in urine by liquid chromatography – tandem mass spectrometry with polarity switching electrospray ionization. Forensic Science International 221, 57–64. 10.1016/j.forsciint.2012.03.025

Landrigan, P.J., Goldman, L.R., 2011. Children’s vulnerability to toxic chemicals: A challenge and opportunity to strengthen health and environmental policy. Health Affairs 30, 842–850. 10.1377/hlthaff.2011.0151

Li, H. ru, Fu, X. hang, Song, L. ling, Cen, M. qiu, Wu, J., 2023. Association between pyrethroid exposure and risk of depressive symptoms in the general US adults. Environmental Science and Pollution Research 30, 685–698. 10.1007/s11356-022-22203-9

London, L., Flisher, A.J., Wesseling, C., Mergler, D., Kromhout, H., 2005. Suicide and exposure to organophosphate insecticides: Cause or effect? American Journal of Industrial Medicine 47, 308–321. 10.1002/ajim.20147

Mehta, P., Parajuli, R.P., Chronister, B.N.C., Yang, K., Barr, D.B., Tu, X.M., Lopez-Paredes, D., Suarez-Lopez, J.R., 2025. Pesticide and Liver Biomarkers Among Ecuadorian Adolescents and Adults Living in Agricultural Settings. Toxics 13, 685. 10.3390/toxics13080685

Meyer, A., Koifman, S., Koifman, R.J., Moreira, J.C., de Rezende Chrisman, J., Abreu-Villaça, Y., 2010. Mood Disorders Hospitalizations, Suicide Attempts, and Suicide Mortality Among Agricultural Workers and Residents in an Area With Intensive Use of Pesticides in Brazil. Journal of Toxicology and Environmental Health, Part A 73, 866–877. 10.1080/15287391003744781

Miyazawa, A., Fujiyoshi, Y., Unwin, N., 2003. Structure and gating mechanism of the acetylcholine receptor pore.

Moreno-Montero, E., Moreta-Herrera, R., Rodas, J.A., Mayorga-Lascano, M., Ruiz, Á., Játiva, R., 2024. Examining the psychometric properties of the Generalized Anxiety Disorder Scale (GAD7) among Ecuadorian university students. Revista de Psicopatologia y Psicologia Clinica 29, 249–260. 10.5944/rppc.39064

Muñoz-Navarro, R., Cano-Vindel, A., Moriana, J.A., Medrano, L.A., Ruiz-Rodríguez, P., Agüero-Gento, L., Rodríguez-Enríquez, M., Pizà, M.R., Ramírez-Manent, J.I., 2017. Screening for generalized anxiety disorder in Spanish primary care centers with the GAD-7. Psychiatry Research 256, 312–317. 10.1016/j.psychres.2017.06.023

Naughton, S.X., Terry, A.V., 2018. Neurotoxicity in acute and repeated organophosphate exposure. Toxicology 408, 101–112. 10.1016/j.tox.2018.08.011

Parajuli, R.P., Chronister, B.N.C., Barr, D.B., Suárez-López, J.R., 2025. Urinary pesticide biomarkers from adolescence to young adulthood in an agricultural setting in Ecuador: Study of secondary exposure to pesticides among children, adolescents, and adults (ESPINA) 2016 and 2022 examination data. Data in Brief 61, 111882. 10.1016/j.dib.2025.111882

Pearson Canada Assessment, 2020. Multidimensional Anxiety Scale for Children 2nd EditionTM (MASC 2TM). Pearson Canada Assessment, Inc.

Picciotto, M.R., Lewis, A.S., Van Schalkwyk, G.I., Mineur, Y.S., 2015. Mood and anxiety regulation by nicotinic acetylcholine receptors: A potential pathway to modulate aggression and related behavioral states. Neuropharmacology 96, 235–243. 10.1016/j.neuropharm.2014.12.028

Rasmussen, A.R., Wohlfahrt-Veje, C., De Renzy-Martin, K.T., Hagen, C.P., Tinggaard, J., Mouritsen, A., Mieritz, M.G., Main, K.M., 2015. Validity of self-assessment of pubertal maturation. Pediatrics 135, 86–93. 10.1542/peds.2014-0793

Roberts, J.R., Karr, C.J., Council On Environmental Health, 2012. Pesticide exposure in children. Pediatrics 130, e1765–1788. 10.1542/peds.2012-2758

Saeedi Saravi, Seyed Soheil, Amirkhanloo, R., Arefidoust, A., Yaftian, R., Saeedi Saravi, Seyed Sobhan, Shokrzadeh, M., Dehpour, A.R., 2016. On the effect of minocycline on the depressive-like behavior of mice repeatedly exposed to malathion: interaction between nitric oxide and cholinergic system. Metabolic Brain Disease 31, 549–561. 10.1007/s11011-015-9764-z

Sánchez-Villena, A.R., Farfán Cedrón, E., 2019. Análisis Factorial Exploratorio del Inventario de Depresión de Beck (BDI-II) en Universitarios Cajamarquinos. Interacciones 9–9.

Sanne, B., Mykletun, A., Moen, B.E., Dahl, A.A., Tell, G.S., 2004. Farmers are at risk for anxiety and depression: The Hordaland Health Study. Occupational Medicine 54, 92–100. 10.1093/occmed/kqh007

Sansone, R.A., Sansone, L.A., 2010. Psychiatric disorders: A global look at facts and figures. Psychiatry (Edgemont) 7, 16–19.

Simcox, N.J., Fenske, R.A., Wolz, S.A., Lee, I.C., Kalman, D.A., 1995. Pesticides in household dust and soil: Exposure pathways for children of agricultural families. Environmental Health Perspectives 103, 1126–1134. 10.1289/ehp.951031126

Simon-Delso, N., Amaral-Rogers, V., Belzunces, L.P., Bonmatin, J.M., Chagnon, M., Downs, C., Furlan, L., Gibbons, D.W., Giorio, C., Girolami, V., Goulson, D., Kreutzweiser, D.P., Krupke, C.H., Liess, M., Long, E., Mcfield, M., Mineau, P., Mitchell, E.A., Morrissey, C.A., Noome, D.A., Pisa, L., Settele, J., Stark, J.D., Tapparo, A., Van Dyck, H., Van Praagh, J., Van Der Sluijs, J.P., Whitehorn, P.R., Wiemers, M., 2015. Systemic insecticides (Neonicotinoids and fipronil): Trends, uses, mode of action and metabolites. Environmental Science and Pollution Research 22, 5–34. 10.1007/s11356-014-3470-y

Siteneski, A., Gómez Mieles, V.S., Romero Riaño, P.A., Montes Escobar, K., Lapo-Talledo, G.J., Dueñas-Rodriguez, A.V., Palma Cedeño, M.A., Villacis Lascano, Y.C., Echeverria Zurita, L.O., 2024. High levels of anxiety and depression in women farmers from Ecuador: A cross-section study in Coastal and Highlands regions. International Journal of Social Psychiatry. 10.1177/00207640241260017

Sparks, T.C., Watson, G.B., Loso, M.R., Geng, C., Babcock, J.M., Thomas, J.D., 2013. Sulfoxaflor and the sulfoximine insecticides: Chemistry, mode of action and basis for efficacy on resistant insects. Pesticide Biochemistry and Physiology 107, 1–7. 10.1016/j.pestbp.2013.05.014

Spitzer, R.L., Kroenke, K., Williams, J.B.W., Löwe, B., 2006. A Brief Measure for Assessing Generalized Anxiety Disorder The GAD-7. Archives of internal medicine 166, 1092–1097.

Suarez-Lopez, J.R., Checkoway, H., Jacobs, D.R.J., Al-Delaimy, W.K., Gahagan, S., Jose R. Suarez-Lopez, Harvey Checkoway, David R. Jacobs Jr, Wael K. Al-Delaimy, Bender, S.G., Suarez-Lopez, J.R., Checkoway, H., Jacobs, D.R.J., Al-Delaimy, W.K., Gahagan, S., Jose R. Suarez-Lopez, Harvey Checkoway, David R. Jacobs Jr, Wael K. Al-Delaimy, Bender, S.G., Suarez-Lopez, J.R., Checkoway, H., Jacobs, D.R.J., Al-Delaimy, W.K., Gahagan, S., 2017. Potential short-term neurobehavioral alterations in children associated with a peak pesticide spray season: The Mother’s Day flower harvest in Ecuador. Neurotoxicology 60, 125–133. 10.1016/j.neuro.2017.02.002

Suarez-Lopez, J.R., Hood, N., Suárez-Torres, J., Gahagan, S., Gunnar, M.R., López-Paredes, D., 2019. Associations of acetylcholinesterase activity with depression and anxiety symptoms among adolescents growing up near pesticide spray sites. International Journal of Hygiene and Environmental Health 222, 981–990. 10.1016/j.ijheh.2019.06.001

Suarez-Lopez, J.R., Jacobs, D.R.J., Himes, J.H., Alexander, B.H., 2013. Acetylcholinesterase activity, cohabitation with floricultural workers, and blood pressure in Ecuadorian children. Environmental health perspectives 121, 619–624. 10.1289/ehp.1205431

Suarez-Lopez, J.R., Jacobs, D.R. Jr., Himes, J.H., Alexander, B.H., Lazovich, D., Gunnar, M., Gunnar, Gunnar, M., Gunnar, Gunnar, M., Gunnar, Gunnar, M., 2012. Lower acetylcholinesterase activity among children living with flower plantation workers. Environmental Research 114, 53–59. 10.1016/j.envres.2012.01.007

Suarez-Lopez, J.R., Lopez, S., Nguyen, A., Klas, J., Gahagan, S., Checkoway, H., Lopez, D., David, P., Jr, R.J., Noble, M., 2020. Associations of Acetylcholinesterase Inhibition Between Pesticide Spray Seasons with Depression and Anxiety Symptoms in Adolescents, and the Role of Sex and Adrenal Hormones on Gender Moderation. Exposure and Health. 10.1007/s12403-020-00361-w

Suk, W.A., Ahanchian, H., Asante, K.A., Carpenter, D.O., Diaz-Barriga, F., Ha, E.H., Huo, X., King, M., Ruchirawat, M., da Silva, E.R., Sly, L., Sly, P.D., Stein, R.T., van den Berg, M., Zar, H., Landrigan, P.J., 2016. Environmental pollution: An underrecognized threat to children’s health, especially in low-and middle-income countries. Environmental Health Perspectives 124, A43–A45. 10.1289/ehp.1510517

Vázquez Morejón, A.J., Vázquez-Morejón Jiménez, R., Zanin, G.B., 2014. Beck Anxiety Inventory: psychometric characteristics in a sample from the clinical Spanish population. Span J Psychol 17, E76. 10.1017/sjp.2014.76

Vijverberg, H.P.M., vanden Bercken, J., 1990. Neurotoxicological effects and the mode of action of pyrethroid insecticides. Critical Reviews in Toxicology 21, 105–126. 10.3109/10408449009089875

Wambua, D., Roman, W., Vidanage, I., Vidal, M., Calafat, A.M., Ospina, M., 2023. Online solid phase extraction high-performance liquid chromatography – Isotope dilution – Tandem mass spectrometry quantification of organophosphate pesticides, synthetic pyrethroids, and selected herbicide metabolites in human urine. Chemosphere 340, 139863. 10.1016/j.chemosphere.2023.139863

Watson, G.B., Loso, M.R., Babcock, J.M., Hasler, J.M., Letherer, T.J., Young, C.D., Zhu, Y., Casida, J.E., Sparks, T.C., 2011. Novel nicotinic action of the sulfoximine insecticide sulfoxaflor. Insect Biochemistry and Molecular Biology 41, 432–439. 10.1016/j.ibmb.2011.01.009

Weisskopf, M.G., Moisan, F., Tzourio, C., Rathouz, P.J., Elbaz, A., 2013. Pesticide exposure and depression among agricultural workers in France. American journal of epidemiology 178, 1051–8. 10.1093/aje/kwt089

Wesseling, C., van Wendel de Joode, B., Keifer, M., London, L., Mergler, D., Stallones, L., 2010. Symptoms of psychological distress and suicidal ideation among banana workers with a history of poisoning by organophosphate or n-methyl carbamate pesticides. Occupational and environmental medicine 67, 778–84. 10.1136/oem.2009.047266

World Health Organization, 2017. Depression and Other Common Mental Disorders. Global Health Estimates. World Health Organization, 2008. 2008 Training Course on Child Grown Assessment 7, 25–36.

Zhang, D., Lu, S., 2022. Human exposure to neonicotinoids and the associated health risks: A review. Environment International 163. 10.1016/j.envint.2022.107201

