## Supplemental Tables 1-3 and Supplemental Figure 1 for "Urinary Pesticide Biomarker Associations with Depression and Anxiety Scores from Adolescence into Young Adulthood in the ESPINA cohort"

Supplemental Table 1. Metabolite concentrations (ug/L) among adolescents of the ESPINA 2016 examination (FUY-8b; n=510, 12–17-year-olds) (Wet Weights).

| Chemical class | Metabolite Name | Acronym | Parent chemical | LOD | Interfering Substance (n) | % Detectable | Geometric Mean (95% CI) | Percentile (95% CI) | | | | | N |
|---|---|---|---|---|---|---|---|---|---|---|---|---|---|
| | | | | | | | | 25% | 50% | 75% | 90% | 95% | |
| Organophosphate | para-Nitrophenol | PNP | Parathion, Methyl parathion | 0.1 | 5.0 | 100.00% | 0.52 (0.49, 0.54) | 0.36 (0.34, 0.38) | 0.50 (0.47, 0.53) | 0.73 (0.67, 0.78) | 1.13 (0.96, 1.29) | 1.46 (1.27, 1.66) | 505 |
| | 3,5,6-Trichloro-2-pyridinol | TCPy | Chlorpyrifos, Chlorpyrifos-methyl | 0.1 | 0.0 | 100.00% | 2.67 (2.49, 2.86) | 1.55 (1.41, 1.68) | 2.67 (2.47, 2.86) | 4.46 (4.03, 4.88) | 6.97 (6.22, 7.72) | 9.49 (7.47,11.51) | 510 |
| | 2-isopropyl-4-methyl-6-hydroxypyrimidine | IMPY | Diazinon | 0.1 | 3.0 | 23.81% | * | <LOD | <LOD | <LOD | 0.29 (0.22, 0.36) | 0.55 (0.36, 0.73) | 507 |
| | Malathion dicarboxylic acid | MDA | Malathion | 0.5 | 0.0 | 37.69% | 0.51 (0.48, 0.53) | <LOD | <LOD | 0.77 (0.74, 0.79) | 0.93 (0.88, 0.98) | 1.06 (0.92, 1.20) | 510 |
| Pyrethroid | 3-phenoxybenzoic acid | 3-PBA | Cyhalothrin, Cypermethrin, Deltamethrin, Fenpropathrin, Permethrin, Tralomethrin | 0.1 | 26.0 | 88.65% | 0.53 (0.50, 0.55) | 0.42 (0.22, 0.63) | 0.42 (0.22, 0.63) | 0.42 (0.22, 0.63) | 0.99 (0.68, 1.31) | 1.95 (1.14, 2.76) | 486 |
| | 4-fluoro-3-phenoxybenzoic acid | 4F-PBA | Cyfluthrin, Flumethrin | 0.1 | 0.0 | 0.00% | * | <LOD | <LOD | <LOD | <LOD | <LOD | 505 |
| | trans-3-(2,2-Dichlorovinyl)-2,2-dimethylcyclopropane carboxylic acid | trans-DCCA | Permethrin; Cypermethrin; Cyfluthrin | 0.6 | 12.0 | 16.09% | * | <LOD | <LOD | <LOD | 0.99 (0.68, 1.31) | 1.95 (1.14, 2.76) | 498 |
| Neonicotinoids | 5-Hydroxy imidacloprid | OHIM | Imidacloprid | 0.4 | 41.0 | 28.54% | * | <LOD | <LOD | 0.45 (0.30, 0.61) | 1.07 (0.88, 1.26) | 1.39 (1.00, 1.77) | 471 |
| | Acetamiprid-N-desmethyl | AND | Acetamiprid | 0.2 | 18.0 | 37.84% | 0.26 (0.24, 0.29) | <LOD | <LOD | 0.44 (0.35, 0.53) | 0.98 (0.67, 1.30) | 2.35 (1.61, 3.10) | 492 |
| | Imidacloprid | IMID | Imidacloprid | 0.4 | 10.0 | 7.72% | * | <LOD | <LOD | <LOD | <LOD | 0.48 (0.42, 0.54) | 501 |
| | Clothianidin | CLOT | Clothianidin, Thiamethoxam | 0.2 | 0.0 | 2.84% | * | <LOD | <LOD | <LOD | <LOD | <LOD | 510 |
| | Acetamiprid | ACET | Acetamiprid | 0.3 | 1.0 | 1.14% | * | <LOD | <LOD | <LOD | <LOD | <LOD | 509 |
| | Thiacloprid | THIA | Thiacloprid | 0.03 | 0.0 | 0.00% | * | <LOD | <LOD | <LOD | <LOD | <LOD | 505 |

LOD= limit of detection.
Values below LOD were assigned LOD/sqrt(2).
Values presented are Geometric Mean or percentile (95% confidence interval).
Ns were calculated by subtracting observations with interfering substances from measured samples.
*If the proportion of detectable concentrations were below 30%, geometric means were not calculated.

Supplemental Table 2. Metabolite concentrations (ug/L) among adolescents and adults of the ESPINA 2022 examination (FUY-14a; n=485, 17–23-year-olds) (Wet Weights).

| Chemical class | Metabolite Name | Acronym | Parent chemical | LOD | Interfering Substance (n) | % Detectable | Geometric Mean (95% CI) | Percentile (95% CI) | | | | | N |
|---|---|---|---|---|---|---|---|---|---|---|---|---|---|
| | | | | | | | | 25% | 50% | 75% | 90% | 95% | |
| Organophosphate | para-Nitrophenol | PNP | Parathion, Methyl parathion | 0.1 | 0 | 100.00% | 0.64 (0.61, 0.68) | 0.41 (0.38, 0.44) | 0.63 (0.59, 0.67) | 0.98 (0.94, 1.03) | 1.37 (1.25, 1.48) | 1.62 (1.46, 1.77) | 485 |
| | 3,5,6-Trichloro-2-pyridinol | TCPy | Chlorpyrifos, Chlorpyrifos-methyl | 0.1 | 0 | 100.00% | 3.44 (3.21, 3.69) | 2.04 (1.80, 2.27) | 3.38 (3.06, 3.70) | 5.68 (5.14, 6.21) | 8.98 (8.03, 9.92) | 12.94 (9.86,16.02) | 485 |
| | 2-isopropyl-4-methyl-6-hydroxypyrimidine | IMPY | Diazinon | 0.1 | 0 | 28.87% | * | <LOD | <LOD | 0.12 (0.08, 0.16) | 0.43 (0.29, 0.56) | 0.85 (0.65, 1.05) | 485 |
| | Malathion dicarboxylic acid | MDA | Malathion | 0.5 | 0 | 9.28% | * | <LOD | <LOD | <LOD | <LOD | 0.69 (0.57, 0.81) | 485 |
| Pyrethroid | 3-phenoxybenzoic acid | 3-PBA | Cyhalothrin, Cypermethrin, Deltamethrin, Fenpropathrin, Permethrin, Tralomethrin | 0.1 | 1 | 98.76% | 0.52 (0.48, 0.56) | 0.31 (0.28, 0.33) | 0.50 (0.46, 0.54) | 0.86 (0.77, 0.95) | 1.39 (1.17, 1.62) | 1.94 (1.45, 2.43) | 484 |
| | 4-fluoro-3-phenoxybenzoic acid | 4F-PBA | Cyfluthrin, Flumethrin | 0.5 | 0 | 0.00% | * | <LOD | <LOD | <LOD | <LOD | <LOD | 485 |
| | trans-3-(2,2-Dichlorovinyl)-2,2-dimethylcyclopropane carboxylic acid | *trans*-DCCA | Permethrin; Cypermethrin; Cyfluthrin | 0.6 | 0 | 42.89% | 0.66 (0.62, 0.70) | <LOD | <LOD | 0.93 (0.84, 1.02) | 1.58 (1.37, 1.78) | 2.46 (1.82, 3.10) | 485 |
| Neonicotinoid | 5-Hydroxy imidacloprid | OHIM | Imidacloprid | 0.10 | 28 | 92.78% | 0.55 (0.50, 0.62) | 0.24 (0.21, 0.28) | 0.50 (0.42, 0.58) | 1.22 (1.07, 1.37) | 2.44 (1.91, 2.97) | 4.17 (2.98, 5.37) | 457 |
| | Acetamiprid-N-desmethyl | AND | Acetamiprid | 0.15 | 10 | 95.16% | 0.35 (0.32, 0.39) | 0.15 (0.13, 0.17) | 0.32 (0.28, 0.36) | 0.66 (0.53, 0.79) | 1.79 (1.43, 2.15) | 2.90 (2.18, 3.61) | 475 |
| | Clothianidin | CLOT | Clothianidin, Thiamethoxam | 0.1 | 148 | 74.18% | 0.34 (0.30, 0.38) | 0.07 (0.01, 0.13) | 0.38 (0.32, 0.45) | 0.84 (0.74, 0.94) | 1.34 (1.16, 1.53) | 1.69 (1.45, 1.93) | 337 |
| Sulfoximine | Sulfoxaflor Isomer | SLF1 | Sulfoxaflor | 0.05 | 11 | 37.55% | 0.07 (0.06, 0.08) | <LOD | <LOD | 0.11 (0.08, 0.13) | 0.31 (0.15, 0.48) | 0.88 (0.62, 1.14) | 474 |
| | Sulfoxaflor Isomer 2 | SLF2 | Sulfoxaflor | 0.05 | 1 | 36.98% | 0.07 (0.06, 0.07) | <LOD | <LOD | 0.10 (0.07, 0.13) | 0.33 (0.18, 0.48) | 0.77 (0.59, 0.95) | 484 |

LOD= limit of detection.
Values below LOD were assigned LOD/sqrt(2).
Values presented are Geometric Mean or percentile (95% confidence interval).
Ns were calculated by subtracting observations with interfering substances from measured samples.
*If the proportion of detectable concentrations were below 30%, geometric means were not calculated.

Supplemental Table 3. Quantile g-computation estimates per quartile increase in the pesticide mixture in relation to depression and anxiety scores (Ψ [95% CI]), or odds ratio of having elevated depression and anxiety (OR [95% CI]), cross-sectionally and longitudinally.

| | **Change in anxiety or depression score for one quartile increase in weighted quantile sum regression index Ψ (95% CI)** | | | | | |
|---|---|---|---|---|---|---|
| | **2016 Assessment** | | **2022 Assessment** | | **Longitudinal Associations** | |
| **Mixture Group** | **Depression CDI-2 Score** | **Anxiety MASC-2 Score** | **Depression BDI-II Score** | **Anxiety GAD-7 Score** | **Depression z-score** | **Anxiety z-score** |
| **Overall Pesticide Mixture** | -2.23 (-5.33, 0.88), 0.16 | 0.18 (-3.00, 3.36), 0.91 | -1.41 (-4.74, 1.93), 0.41 | -0.50 (-1.94, 0.95), 0.50 | -0.02 (-0.17, 0.12), 0.74 | 0.02 (-0.13, 0.16), 0.83 |
| **Organophosphate Mixture** | -1.75 (-93.96, 90.46), 0.97 | 0.25 (-2.01, 2.52), 0.83 | 0.18 (-0.79, 1.15), 0.72 | 0.24 (-0.18, 0.66), 0.25 | 0.004 (-0.08, 0.09), 0.92 | 0.001 (-0.08, 0.09), 0.98 |
| **Pyrethroid Mixture** | - | - | -1.06 (-3.05, 0.94), 0.30 | -0.64 (-1.50, 0.22), 0.15 | -0.05 (-0.19, 0.08), 0.43 | -0.01 (-0.12, 0.10), 0.89 |
| **Neonicotinoid Mixture** | - | - | 0.08 (-1.46, 1.62), 0.92 | 0.38 (-0.30), 1.05), 0.27 | -0.02 (-0.13, 0.08), 0.65 | -0.01 (-0.09, 0.10), 0.87 |
| **Sulfoximine Mixture** | - | - | **-3.01 (-5.01, 1.01), 0.003** | **-0.99 (-1.86, -0.14), 0.02** | - | - |
| **Neonicotinoid and Sulfoximine Mixture** | - | - | -0.65 (-1.79, 0.50), 0.27 | -0.78 (-1.85, 0.28), 0.15 | - | - |
| | **Odds of having elevated depression or anxiety scores, compared to those with lesser symptoms, for one quartile increase in the pesticide mixture (OR [95% CI])** | | | | | |
| **Mixture Group** | **Elevated Depression Score (CDI-2≥65)** | **Elevated Anxiety Score (MASC-2≥60)** | **Elevated Depression Score (BDI-2≥20)** | **Elevated Anxiety Score (GAD-7≥5)** | **Elevated Depression Score (CDI-2≥65 or BDI-2≥20)** | **Elevated Anxiety Score (MASC-2≥60 or GAD-7≥5)** |
| **Overall Pesticide Mixture** | 0.92 (0.57, 1.48), 0.74 | 1.05 (0.80, 1.39), 0.72 | 0.70 (0.41, 1.18), 0.18 | 1.09 (0.78, 1.52), 0.62 | 1.10 (0.91, 1.33), 0.31 | 1.04 (0.98, 1.12), 0.20 |
| **Organophosphate Mixture** | 0.92 (0.64, 1.32), 0.64 | 1.04 (0.85, 1.27), 0.68 | 1.01 (0.89, 1.15), 0.85 | 1.02 (0.93, 1.13), 0.66 | 1.06 (0.97, 1.15), 0.23 | 1.00 (0.96, 1.04), 0.96 |
| **Pyrethroid Mixture** | - | - | 1.02 (0.85, 1.23), 0.80 | 0.93 (0.80, 1.07), 0.32 | 1.04 (0.90, 1.21), 0.59 | 1.03 (0.98, 1.09), 0.23 |
| **Neonicotinoid Mixture** | - | - | 1.03 (0.85, 1.24), 0.77 | **1.21 (1.05, 1.40), 0.01** | 1.04 (0.93, 1.17), 0.49 | 1.02 (0.98, 1.07), 0.88 |
| **Sulfoximine Mixture** | - | - | **0.54 (0.38, 0.77), 0.001** | 0.95 (0.79, 1.15), 0.62 | - | - |
| **Neonicotinoid and Sulfoximine Mixture** | - | - | 0.67 (0.43, 0.17), 0.08 | 1.14 (0.88, 1.49), 0.32 | - | - |

2016 models were adjusted for age, gender, race, tanner, parental education, BMI-for-age z-score, and ln-creatinine.
2022 models were adjusted for age, gender, race, specific gravity, BMI, monthly household income, and education level.
Longitudinal models were adjusted for age, gender, race, specific gravity, BMI, monthly household income, and education level.

Mixture Groups included the following metabolites for the varying analyses.
2016 Analyses:
Overall Pesticide Mixture: PNP, TCPy, MDA, 3-PBA, and AND.
Organophosphate Mixture: PNP, TCPy, and MDA.
As only 1 metabolite was above the detection threshold of 38%, we did not run mixture analyses for the pyrethroids nor neonicotinoids for this time point.

2022 Analyses
Overall Pesticide Mixture: PNP, TCPy, 3-PBA, *trans*-DCCA, OHIM, AND, CLOT, SLF1, and SLF2.

Organophosphate Mixture: PNP and TCPy.
Pyrethroid Mixture:3-PBA and trans-DCCA.
Neonicotinoid Mixture: AND, OHIM, and CLOT
Sulfoximine Mixture: SLF1 and SLF2
Neonicotinoid and Sulfoximine Mixture: AND, OHIM, CLOT, SLF1 and SLF2

Longitudinal Analyses
Overall Pesticide Mixture: PNP, TCPy, 3-PBA, trans-DCCA, AND, and OHIM.
Organophosphate Mixture: PNP and TCPy
Pyrethroid Mixture: *trans*-DCCA and 3-PBA
Neonicotinoid Mixture: OHIM and AND

Supplemental Figure 1. Cross-sectional quantile g-computation positive and negative weights for the overall pesticide mixture, neonicotinoid mixture, and sulfoximine mixture with elevated depression and anxiety symptoms.

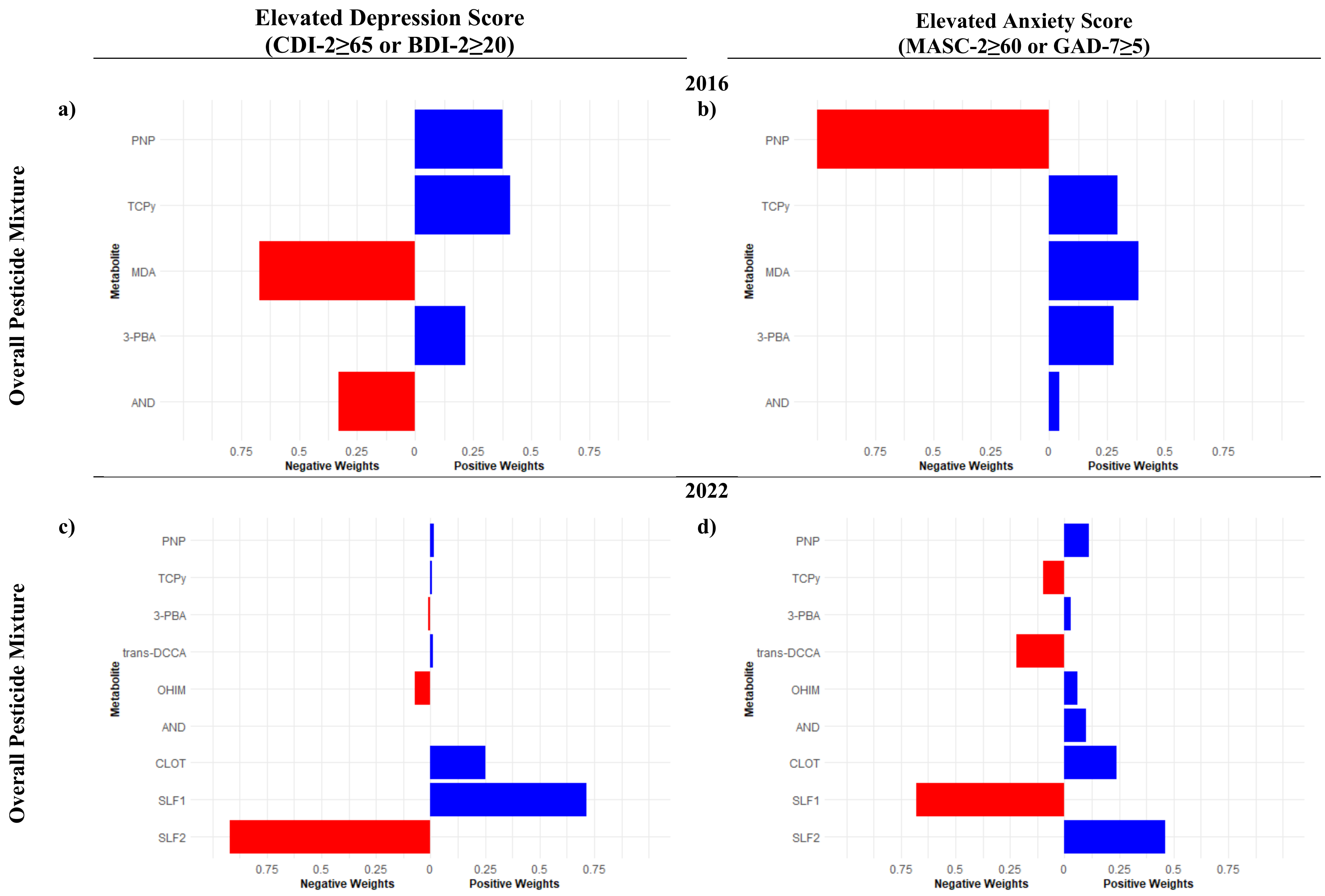

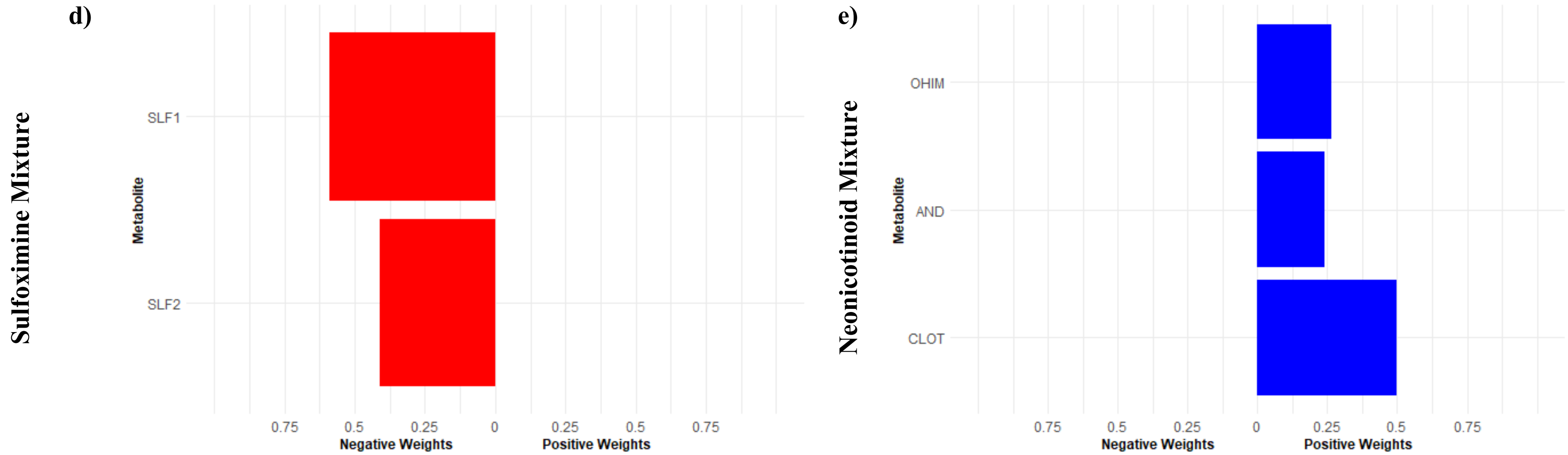


Mixture Groups included the following metabolites for the varying analyses.

2016 Analyses:

Overall Pesticide Mixture: PNP, TCPy, MDA, 3-PBA, and AND.

2022 Analyses

Neonicotinoid Mixture: AND, OHIM, and CLOT

Sulfoximine Mixture: SLF1 and SLF2
